# A MULTI-MINERAL INTERVENTION TO IMPROVE DISEASE-RELATED AND MECHANISTIC BIOMARKERS IN ULCERATIVE COLITIS PATIENTS

**DOI:** 10.1101/2025.02.17.25322298

**Authors:** Muhammad N. Aslam, Danielle (Kim) Turgeon, Henry D. Appelman, Ryan Stidham, Shannon McClintock, Ron Allen, Gillian Moraga, Isabelle Harber, Kara J. Jencks, Molly M. McNeely, Ananda Sen, Karl J. Jepsen, James Varani

## Abstract

**Introduction:** The long-term goal of our studies is to determine if, and to what extent, a multi-mineral product (Aquamin) could have beneficial impact on individuals with ulcerative colitis (UC). As a step toward achieving that goal, we carried out a 180-day biomarker trial in patients with UC in remission or at the mild stage.

**Approach:** A total of 28 subjects were included in the study. Each was randomized to receive either Aquamin for 180 days or placebo for the first 90 days. At day-90, placebo subjects crossed over to Aquamin for the final 90 days. At days-0, −90 and −180, serum samples were assessed for alkaline phosphatase (ALP), intestine-specific ALP (ALPI), C-reactive protein (CRP) and for biomarkers of bone turnover (osteocalcin, TRAP5b and bone-specific ALP [e.g., BALP]). Stool specimens were assessed for fecal calprotectin at the same time points and colon biopsies were examined histologically. Each subject underwent DEXA scanning (day-0 and −180 only). In addition, a mass spectrometry-based proteomic assessment was performed using colon biopsy specimens obtained at each time point.

**Results:** Subjects receiving Aquamin for the complete 180-day period (a total of 12) demonstrated improvement in all biomarkers; this was not seen in the placebo group (16 subjects). Subjects who received Aquamin for 90-days were intermediary in their responses. Subjects receiving Aquamin for 180-days also demonstrated increases in bone mineral density (BMD) and bone mineral content (BMC) resulting in a statistically-significant increase in the hip strength index over the period of treatment. This was accompanied by increases in osteocalcin and TRAP5b and by a decrease in BALP. The proteomic screen demonstrated up-regulation of multiple gut barrier proteins, cell surface transporter molecules and certain proteins with anti-inflammatory potential in response to Aquamin. Aquamin treatment also led to down-regulation of several proteins associated with the pro-inflammatory state.

**Conclusion:** These findings suggest the potential value of multi-mineral intervention (Aquamin) as a low-cost, non-toxic adjuvant therapy for mild UC or for individuals with UC in remission.

## INTRODUCTION

Ulcerative colitis (UC) is a chronic disease manifested as diffuse mucosal inflammation along with superficial ulcer formation in the inner lining of the large intestine (1,2). Large bowel injury may be focal in nature or widespread. Additional extra-colonic features may also exist; among the most prevalent are liver inflammation (especially of the bile duct) (3,4) and a propensity toward bone loss (5). Dysregulation of the immune system (1,2) is thought to be the underlying pathophysiological mechanism, and the therapeutic armamentarium currently used for UC is aimed, primarily, at controlling inflammation (6). Agents that target inflammation broadly including steroids and non-steroidal anti-inflammatory drugs such as mesalamine have long been treatment options. More recently, biological agents (small molecules and antibodies) targeting specific components of the inflammatory process have become part of the treatment combination (7). The use of biologicals has dramatically expanded in recent years, but clinical remission rates continue to be approximately 20% over placebo in clinical trials (6), suggesting that additional aspects of UC pathophysiology need to be addressed.

Abnormal barrier function in the large bowel, leading to increased mucosal permeability, is another potentially important pathologic feature of UC (8–11). The colonic barrier is designed to isolate the interstitium from colonic contents (e.g., bacterial cells and cell wall components, soluble bacterial toxins, pollens, other particulates, food allergens) while still providing for selective transport of nutrients and essential minerals across the colonic wall. While inflammation, itself, is a cause of barrier dysfunction (9–11), pre-existing barrier defects provide a milieu in which inflammation can take hold (10). While an intact barrier is essential to colonic health, there are no therapies that directly address barrier defects in UC (11,12) or in other inflammatory conditions of the bowel.

Our own past studies have shown that a marine red algae-derived multi-mineral product (Aquamin) can significantly improve gastrointestinal health in experimental animals when the mineral supplement is provided as a dietary component. In long-term (15-18 month) mouse studies, Aquamin inclusion in the diet significantly reduced the incidence of colon polyp formation (13,14) as well as tumors in the liver (15). A reduction in inflammation throughout the gastrointestinal tract and systemically was associated with these beneficial effects (13,14). Studies by Aviello et al demonstrated Aquamin’s ability to inhibit colitis in the IL10-/- mouse (16). In organoid culture studies carried out with human colon tissue from healthy subjects and ulcerative colitis patients, Aquamin treatment reduced the levels of several proteins that promote inflammation and increased the expression of proteins that counteract the pro-inflammatory response (17–19). The same mineral supplement increased the elaboration of numerous proteins that contribute to barrier structure / function (17–21). Subsequently, a biomarker trial in healthy human subjects revealed that many of the same protein changes seen in organoid culture were also seen in colonic biopsies after 90-days of daily Aquamin ingestion (22). Together, these preclinical findings allow us to suggest that Aquamin may have potential as an ancillary treatment in individuals with UC or other chronic inflammatory bowel conditions.

As a way to begin testing this idea, a small biomarker trial was conducted in subjects with UC in remission or with disease at the mild stage. Subjects in the trial were treated with Aquamin for 180 days or with placebo in a one-way cross-over study (placebo to Aquamin at day-90). Before treatment and at day-90 and day-180, colon tissue biopsies and blood were obtained from each subject and assessed for a variety of clinical and research endpoints that could reflect a response to the intervention. In addition, dual-energy X-ray absorptiometry (DEXA) scans were obtained before treatment and at day-180 as bone loss is commonly seen in UC (5) and the same multi-mineral supplement has been shown to retard bone loss in mice (23–25). The results of this study are described herein.

## MATERIALS AND METHODS

### Intervention

Aquamin is the intervention used in this FDA-approved (IND# 141600) study for UC. Aquamin is a calcium-, magnesium- and multi-trace element-rich product obtained from the calcified fronds of red marine algae of the *Lithothamnion* genus (26). The calcium and magnesium ratio in Aquamin is approximately twelve to one (12:1); Aquamin also contains measurable levels of seventy-two additional trace elements. Aquamin is sold as a dietary supplement (GRAS 000028; (Marigot Ltd, Cork, Ireland) and is used in various products for human consumption in Europe, Asia, Australia, and North America. A single batch of Aquamin-TG^®^ (Food Grade) was used for the trial presented here. It was provided in hydroxypropyl methylcellulose (HPMC) capsules formulated to contain 600 mg of Aquamin standardized to 200 mg of calcium per capsule. Patients were prescribed four capsules of Aquamin (formulated to deliver 800 mg of calcium) per day (two in the morning and two in the evening) in addition to their ongoing UC maintenance medications. Supplement Table 1 provides a complete list of elements detected in Aquamin and their relative amounts taken on a daily basis. Mineral composition of Aquamin-TG^®^ was established via an independent laboratory (Advanced Laboratories; Salt Lake City, Utah) using Inductively Coupled Plasma Optical Emission Spectrometry. Aquamin-TG^®^ has been used in our previous trial (22,27). Maltodextrin was used as a placebo and prescribed in similar capsules with the same weight (600 mg) per capsule.

### Study design, regulatory oversight and study population

This single-site study was a pilot-phase, double-blind, randomized-controlled, one-way cross-over clinical interventional trial in which participants with UC, either in-remission or with mild disease, were included (Study design is shown in Figure 1). The participants were recruited through the Michigan Medicine web portal (UMHealthResearch) and by posting flyers in the hospital. All study interactions and procedures were conducted in the Michigan Clinical Research Unit.

**Figure 1.**
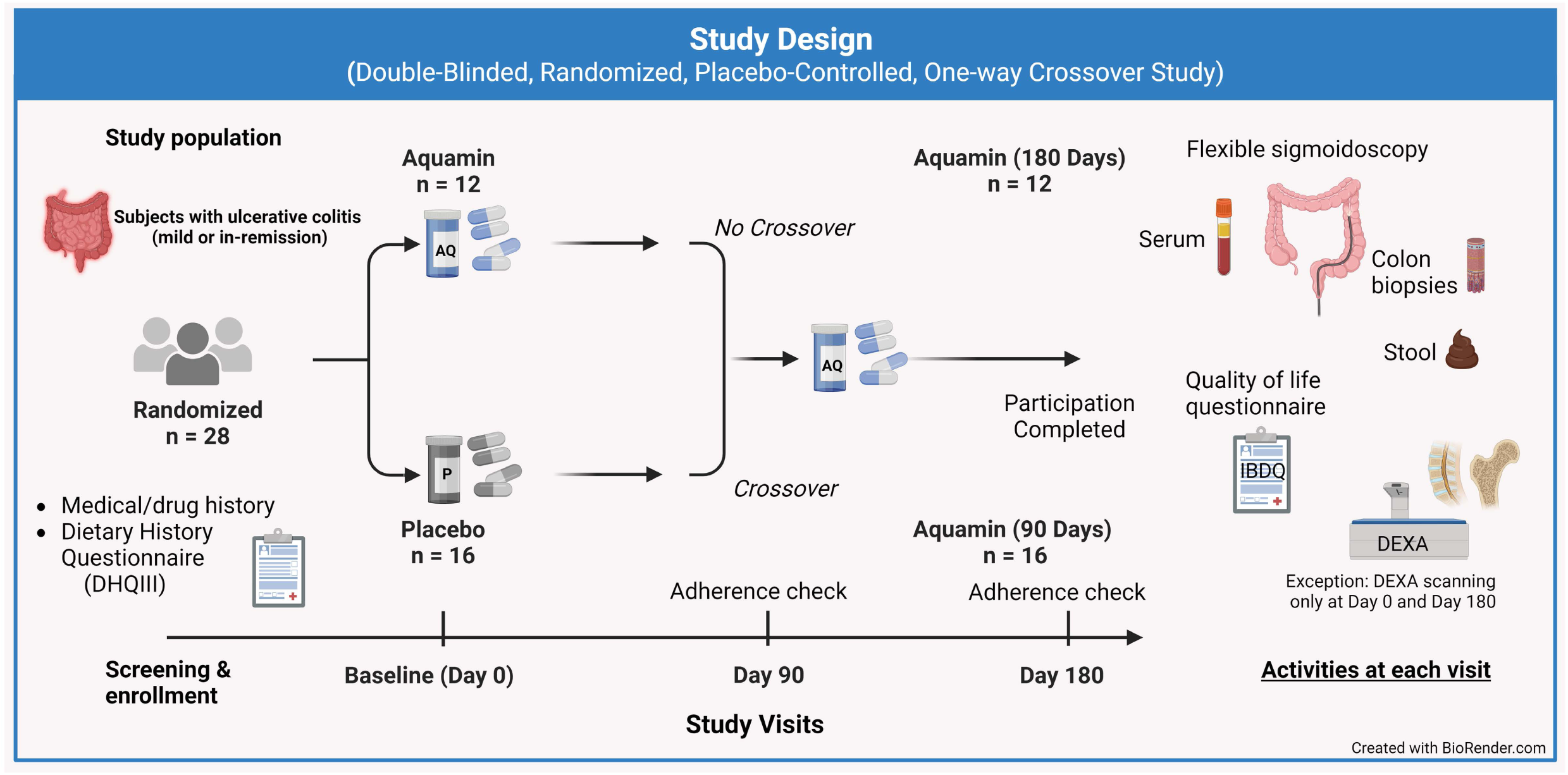
Study design presented in graphical form. Subjects were randomized to either placebo or Aquamin. After 90 days of intervention subjects on placebo crossed over to Aquamin, while subjects on Aquamin remined on Aquamin treatment. There were, thus, three groups: i) Aquamin treatment for 180 days, ii) Placebo for 90 days and iii) Aquamin for 90 days.

The interventional study was conducted with FDA approval of Aquamin as an Investigational New Drug (IND#141600) and with oversight by the Institutional Review Board at the University of Michigan Medical School (IRBMED). The Michigan Institute for Clinical and Health Research (MICHR) provided site monitoring, data collection and data analysis tools (RedCap). A data and safety monitoring committee (DSMC) was responsible for reviewing the study every month to safeguard the welfare of study participants and ensure the study’s appropriateness and data integrity. The study was registered as an interventional clinical trial with details at Clinicaltrials.gov (study identifier NCT03869905). All participants provided written informed consent prior to inclusion. This phase II trial involving human participants was carried out in accordance with recognized ethical guidelines, for example, the Declaration of Helsinki, the International Ethical Guidelines for Biomedical Research Involving Human Subjects (CIOMS), International Council for Harmonisation (ICH) Good Clinical Practice guidelines, the Belmont Report, and the U.S. Common Rule.

A total of 40 subjects were screened and 37 subjects were enrolled in this interventional trial. Four subjects withdrew and five subjects were lost to follow-up. 28 subjects who started on treatment completed the study. This included 12 subjects who received Aquamin for the entire period of 180 days and 16 subjects who were randomized initially to placebo (day-0 to day-90) and crossed over to Aquamin for the final 90-days. Participants were males or non-pregnant females in general good health but having a confirmed diagnosis of UC by histology and endoscopy and were either in-remission or had mild disease at entry with stable maintenance therapy. Exclusion criteria included individuals with severe acute UC, or individuals with Crohn’s disease, gastrointestinal / colonic malignancy or gastrointestinal hemorrhagic disorders. Individuals with coagulopathy or receiving therapeutic doses of Coumadin or heparin were also excluded as were those with a history of kidney disease or kidney stones. A washout period of 30 days was required for certain medications (for example, steroids, antibiotics, non-steroidal anti-inflammatory drugs, or supplements containing calcium or other minerals, Vitamin D, and fiber). [Note: One study participant had completed the initial 90 days of participation immediately prior to the onset of the COVID-19 pandemic. The subject switched over to Aquamin for the last 90 days of the study in accordance with study protocol, but due to the closure of the clinical research unit to make space for COVID-19 emergencies, remained on Aquamin for the complete 180-day period. This subject is included in both the placebo group for the first 90 days and as part of the Aquamin-treatment group for the final phase.]

Figure 1 summarizes the study design of this trial. Each subject had four study visits – i.e., i) screening and enrollment; ii) baseline (day 0), iii) midpoint (day-90) and iv) final visit (day-180). Briefly, at the screening visit, individuals who chose to participate after having the study explained were given the NIH Diet History Questionnaire III (DHQ III), a food frequency questionnaire that includes portion size and dietary supplement questions, as a way to evaluate baseline calcium intake levels (28). Participants were also asked about use of dietary supplements, antibiotics, and non-steroidal anti-inflammatory drugs. A brief drug and medical history was taken and a brief physical examination was given. Supplement Table 2 provides the ongoing therapeutics treatment log of study participants. It also provides disease status at the start of the participation (including pre-participation colonoscopy and pathology report) with year of diagnosis and location and extent of UC lesions. Incidence of the last flare was documented. Subjects also completed a 32 questions-based instrument known as the Inflammatory Bowel Disease Questionnaire (IBDQ) (29). Eligible subjects signed informed consent at the screening visit prior to any study participation.

At the baseline visit, participants underwent flexible sigmoidoscopy (unprepped; *i.e.,* without bowel cleansing procedure). Twelve 2.8 mm colonic biopsies were obtained along with five stool specimens from within the sigmoid colon (20 cm above the anus) using cold Captura Pro biopsy forceps (Cook Medical). If there was evidence of an active UC lesion, the endoscopist avoided these areas while taking biopsies. Tissue samples were saved in 10% formalin for histology or snap frozen in liquid nitrogen and saved at −80°C for proteomic analysis. Stool samples were either transported in liquid nitrogen and stored at −80°C or processed for fecal calprotectin (fCAL) assessment using extraction buffer (BÜHLMANN) and then stored at −20°C. Venous blood samples were taken to run a comprehensive metabolic panel and to assess C-reactive protein (CRP) levels. The panel included total albumin, bilirubin, aspartate aminotransferase (AST), alanine aminotransferase (ALT), and alkaline phosphatase (ALP). The Michigan Medicine laboratory processed these serum samples and generated individual reports for each participant according to their standard operating procedure. Additional serum samples were used to evaluate the intestine-specific form of alkaline phosphatase (ALPI) by ELISA (AssayGenie), and other serum endpoints related to bone health (see below).

After baseline sigmoidoscopy, participants were randomized to either placebo or Aquamin (providing 800 mg of calcium per day). Participants continued with their ongoing maintenance therapy for UC. During the interventional period, participants were contacted by study coordinators monthly to assess study progress and adherence to the study protocol as well as to identify unwanted side effects. Compliance was assessed by capsule log entries and by counting unused capsules returned at the end of each 90-day period.

At the end of the initial 90-day intervention period (90±5 days), participants again underwent unprepped flexible sigmoidoscopy and ten colonic biopsies along with five stool specimens were collected and stored as at baseline. Blood was also taken for serum markers as at baseline. Subjects completed the IBDQ survey again. After the 90-day visit, subjects on Aquamin remained on Aquamin, while subjects on placebo switched over to Aquamin for the final 90 days. At the end of 180-day interventional period, participants underwent the same procedure as at baseline and midpoint and completed all the tasks including capsule count to confirm compliance. [Note: the additional biopsies and stool specimens collected at each timepoint were retained for backup or saved for future use in immunohistochemical, microbial and metabolic analyses. Results from any latter analyses will be reported separately.]

### Fecal calprotectin

Stool samples collected at each visit (day-0, day-90 and day-180) were assessed for fCAL at the end of the study by ELISA (BÜHLMANN). fCAL is a major cytosolic protein of neutrophils and its level in stool specimens is a reliable, quantitative, non-invasive measure of mucosal inflammation in ulcerative colitis (30,31). Previous studies have demonstrated a change in fCAL level corresponding to altered disease activity status. Fresh stool samples were processed after each visit using the extraction buffer (B-CAL-EX) to extract calprotectin and saved at −20°C. ELISAs were conducted at the end of the study to avoid batch-to-batch variability.

### Histological evaluation of colonic biopsies

After formalin-fixation and paraffin-embedding, colon biopsies were sectioned and stained with hematoxylin and eosin (H&E) to assess structural features in the colonic wall and mucosal inflammation. The assessment was performed in a blinded fashion using a modified Geboes Scoring System; this provides a validated semi-quantitative assessment of the histological findings (32). It consists of five grades, ranging from grade 0 (No abnormality) to grade 4 (Ulcer formation). Grades differentiate between quiescent, mildly active, and moderate to severely active disease levels. Higher Geboes scores also provide indication of the severity of mucosal inflammation.

### Symptomatic and Quality of Life UC Assessments

A clinical instrument and quality of life survey – i.e., Inflammatory Bowel Disease Questionnaire (IBDQ) – was used to longitudinally monitor subjects’ UC-related symptoms over the course of the study. It is a 32-item questionnaire that captures disease parameters on four domains of functioning and well-being: bowel and systemic symptoms and emotional and social function (29). The total IBDQ score ranges from 32 to 224 and is generated by tallying up all 32 items. IBDQ was administered on each study visit. Higher IBDQ scores represent improved quality of life. Patients were also asked to share their experience in their own words at the completion of the study or at the close-out call two weeks later.

### DEXA scanning and serum bone biomarkers

DEXA scanning was performed at baseline (Day 0) and the final visit (Day 180) to assess bone changes using a GE Lunar Prodigy ADVANCE Plus (enCORE-based X-ray Bone Densitometer). DEXA outputs provided bone mineral density (BMD), bone mineral content (BMC) and area values for upper femoral shaft including femoral neck, and lumbar spine. These DEXA outputs along with additional variables (buckling ratio, cross-sectional area or moment of inertia and additional geometric parameters) along with variables such as age and gender were used by the software associated with the DEXA scan to calculate a hip strength index as described by Yoshikawa et al (33). DEXA scanning was not done at the day-90 visit with the anticipation that 90 days of intervention would not be long enough to detect differences and would not justify exposure of subjects to the additional radiation. Serum bone turnover markers (Osteocalcin, tartrate-resistant acid phosphatase 5b [TRAP5b] and bone-specific alkaline phosphatase [BALP]) were assessed by ELISAs (AssayGenie) at each time point.

### Proteomic assessment of tissue biopsies – Data-Independent Acquisition

Proteomic assessment of the colon mucosal biopsies collected at each study visit was conducted at the IDeA National Resource for Quantitative Proteomics (Little Rock, AR) using the Data-Independent Acquisition (DIA) approach. A complete description of the technical details for proteomic data acquisition is presented as Supplement Text-File 1.

Following data acquisition, data were searched using an empirically corrected library against the UniProt Homo sapiens database (April 2022) and a quantitative analysis was performed to obtain a comprehensive proteomic profile. Proteins were identified and quantified using EncyclopeDIA (34) and visualized with Scaffold DIA using 1% false discovery thresholds at both the protein and peptide levels. Protein MS2 exclusive intensity values were assessed for quality using ProteiNorm (35). The data were normalized using VSN (36) and analyzed using proteoDA to perform statistical analysis using Linear Models for Microarray Data (limma) with empirical Bayes (eBayes) smoothing to the standard errors (37). The abundance ratio (Log2 fold-change) was calculated by comparing baseline Log2 VSN normalized intensities to the post-intervention intensities. The current study’s proteomic analysis involved a directed search of proteins involved in differentiation, barrier-related cell adhesion proteins, ion transporters and pro- or anti-inflammatory molecules. (Note: The complete set of unbiased findings from the colonic biopsies, will be fully described in a separate report. Additionally, a parallel set of proteomic data obtained from serum samples taken at the same time points and analyzed in the same manner will be included). QIAGEN Ingenuity Pathway Analysis (IPA) was used to find significantly altered pathways by the participating proteins. IPA also provided predictions on the activated or inhibited status of these pathways. This commercial software is based on a knowledge database which can identify specific pathways and it can generate biological networks influenced by a given set of proteins and their observed expression. These proteomics data are deposited to MassIVE (massive.ucsd.edu), a data repository for open access.

### Statistical evaluation

Pre-intervention and post-intervention values were obtained from each subject for the various serum analytes in the metabolic panel, as well as for CRP, fCAL, histological scores, bone DEXA markers and each ELISA marker. Group means and standard deviations were generated and the pre-post data were analyzed by paired t-test to calculate two-tailed p values using 95% confidence level. Then pre-post-intervention values for ALP, ALPI, CRP, fCAL and Geboes scores from the Aquamin-treatment group and placebo group were compared. Group means were used to calculate a composite value for both groups and analyzed for significance by two-tailed unpaired t-test. Similarly, means of serum bone markers (Osteocalcin, TRAP5b and BALP) were combined to obtain a composite score for two-group analysis and significance was assessed by two-tailed unpaired t-test. GraphPad Prism v10.2 was used for these analyses.

Proteomic data were generated by DIA, and the limma software package was used to generate p-values reflective of differences between the means of the two groups (comparator being the respective pre-intervention value). The un-adjusted p-values were corrected using Benjamini-Hochberg method (also known as the False Discovery Rate) in limma. For pathways enrichment analysis, IPA uses the Fisher’s Exact Test to calculate a statistical significance (p-value) of overlap of the dataset molecules with various sets of molecules that represent annotations such as Canonical Pathways. A p-value <0.05 was considered significant. IPA calculates the Z-score by comparing observed expression to the expected expression in the knowledgebase, predicting up or down-regulation and weighted by the underlying findings. A Z-score of ≥ 2 or ≤ −2 is considered significant.

Due to the small sample size, analyses were not adjusted for baseline sociodemographic data including gender, or for existing treatments or baseline dietary calcium intake amounts.

## RESULTS

### Participant characteristics

Twenty-eight subjects completed the study. This included 12 subjects who received Aquamin for 180 days and 16 subjects who were randomized initially to placebo (day-0 to day-90) and crossed over to Aquamin for the final 90-days. Demographic profiles including ethnicity, gender and age are presented in Supplement Table 3. Calcium-intake levels based on the DHQ3 survey recorded at the start of the study and body mass index (BMI) values are also provided in Supplement Table 3. The findings presented in the supplement table allow us to conclude that while subjects were not stratified with regard to demographic characteristics, calcium-intake or BMI, there appeared to be no significant group differences in any of these features that would prevent interpretation of study findings.

### Serum chemistry values and adverse events (AEs)

A panel of serum chemistry values was obtained for each subject prior to the start of the intervention and at day-90 and day-180. The chemistry markers were assessed primarily as part of a safety evaluation but were also used to identify any potentially beneficial response to Aquamin. Results of these studies are presented in Table 1. With the exception of ALP (see below), there was little difference in pre- post-treatment values for the metabolic panel analytes in either treatment group.

Supplement Table 4 provides a list of intervention-related AEs. There were no serious AEs reported and AEs related to gastrointestinal symptoms were similarly distributed between the two cohorts: 29 AEs with placebo and 28 AEs with Aquamin. This is consistent with our previous findings in healthy subjects (27) and suggests that safety or tolerability will not be an issue with Aquamin use for 180 days in UC patients.

### Serum alkaline phosphatase

Among the entire panel of serum markers assessed, only one marker measurably changed over the course of treatment; this was ALP, which decreased by 9.1% (i.e., from 79±19 U/L to 72±16 U/L) in the group of individuals receiving Aquamin for the entire 180-day period (p=0.0046 by paired t-test) (Figure 2A). By comparison, the average ALP level increased by 2.4% in the placebo group (Figure 2A). When placebo and Aquamin groups were compared by two-sample t-test comparing independent means applied on the pre-post differences for ALP, the p-value was equal to 0.07.

**Figure 2.**
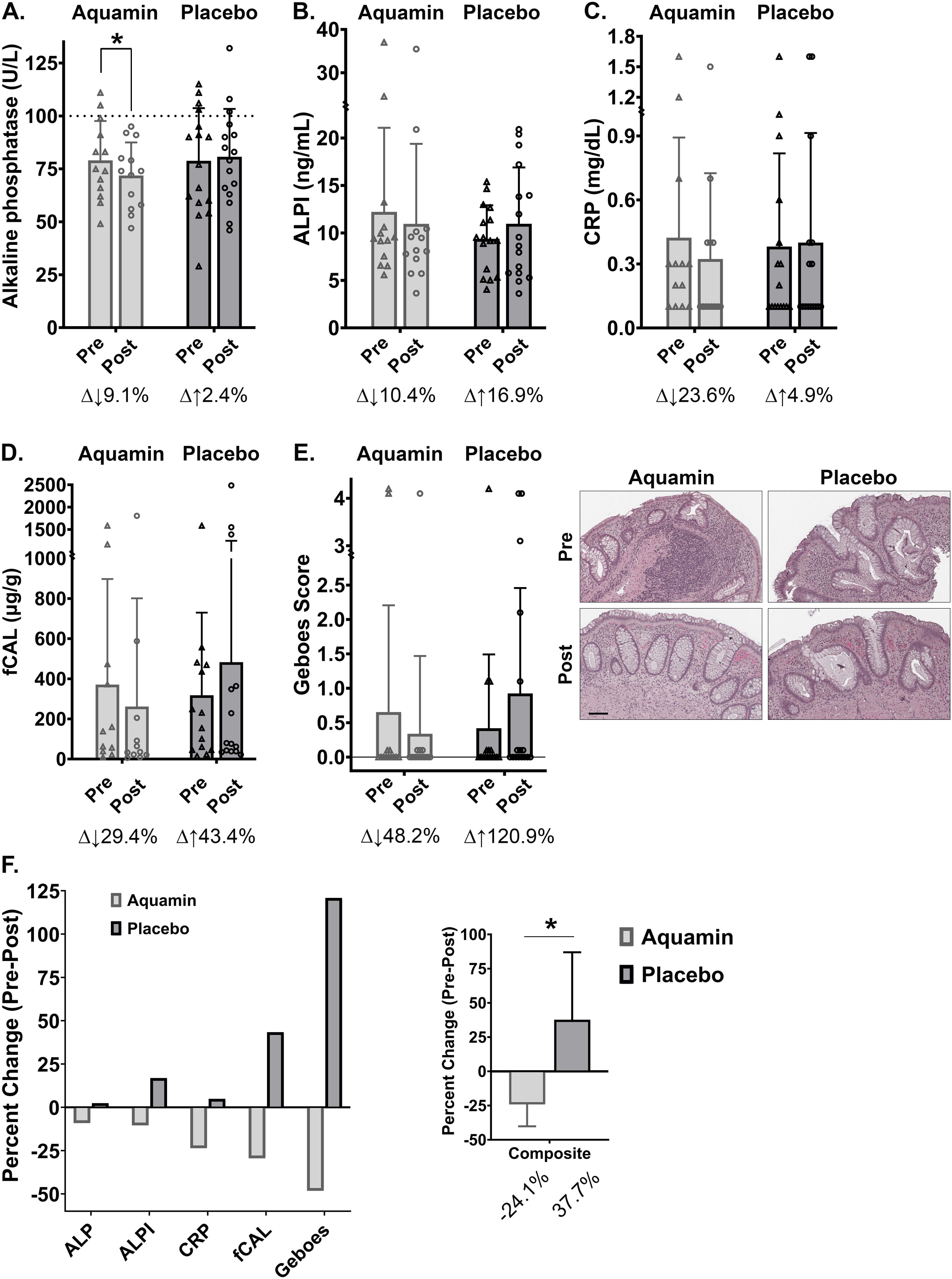
Chemistry and histology endpoints: Aquamin (180-day treatment group) versus placebo. A. Serum alkaline phosphatase (ALP): Values (U/L) determined as part of the serum chemistry panel. **B**. Serum intestine-specific alkaline phosphatase (ALPI): Values (ng/mL) determined by ELISA. **C**. C-reactive protein (CRP): Values (mg/dL) determined in serum. **D**. Fecal calprotectin (fCAL): Values (μg/g) determined by ELISA (BUHLMANN). **E**. Histological assessment: Values presented as a modified Geboes score. **Insert**: Histological appearance of colon tissue from one subject in each treatment group at day-0 and at the end of treatment. All values are treatment group means and standard deviations. Delta values shown beneath each pair of bars indicate percentage change in the post-treatment value relative to the pretreatment value. Statistical significance between pre- and post-treatment values was determined by paired t-test. Asterisk (*) indicates p<0.05. **F.** Aquamin versus placebo comparison: Pre- post-treatment differences for each endpoint are plotted for the two participant groups to show the divergence between the two groups. A composite score was generated by combining the five individual endpoint scores. Statistical significance (p<0.05) between group (composite) values was determined by unpaired t-test.

Serum ALP activity reflects contributions from multiple tissue sources of enzyme (38). As a follow-up, levels of intestine-specific ALP (ALPI) were assessed by ELISA. It can be seen in Figure 2B that the intestinal isoform of ALP decreased by 10.4% in subjects receiving Aquamin over the 180-day treatment period. In contrast, ALPI increased among the 16 individuals in the placebo group by an average of 16.9% (Figure 2B).

When serum ALP data were examined on a subject-by-subject basis, we identified eight individuals whose ALP values were above the 100 U/L level that is the upper limit for normal range (The reference range for normal ALP at the Michigan Medicine Laboratory is 25-100 U/L). Of the eight individuals, three were from the cohort that was randomly assigned to receive Aquamin for 180 days, while the other five were in the placebo group. At the end of the treatment period, there was an average decrease of 12% among the individuals treated with Aquamin (−17%, −10% and −8% compared to pretreatment values; p<0.06 by paired t-test). In contrast, there was virtually no change in the average serum ALP level among the five subjects in the placebo group (19%, 20%, 4%, −7% and −26% compared to pretreatment values). With serum ALPI, there was an average 27% decrease among the three Aquamin-treated subjects (−17%, −23% and −40%, respectively; p<0.05 by paired t-test) while the average ALPI level increased by 26% among the five subjects receiving placebo only (13%, 26%, 43%, 48% and −14% change from pretreatment). This difference in ALPI between the Aquamin-treated and placebo groups was statistically significant (p=0.018 by unpaired t-test).

### Serum C-reactive protein

CRP values are presented in Figure 2C. Prior to initiation of treatment, CRP levels were above baseline in 16 specimens (out of 28 total). Average CRP values were similar between the placebo and Aquamin-treatment groups (0.38 mg/dL and 0.42 mg/dL, respectively). Over the course of treatment, there was a 23.6% decline in CRP values among subjects receiving Aquamin (group average = 0.32 mg/dL) and a 4.9% increase in the placebo group (average = 0.40 mg/dL) (Figure 2C).

### Fecal calprotectin (fCAL) and histological evaluation of colon biopsies

fCAL levels were assessed in stool specimens collected from each subject prior to initiation of treatment and at the end of the treatment period. In parallel, colon biopsies obtained at each time-point were examined histologically using the criteria described in the Materials and Methods Section. As can be seen from Figure 2D, pretreatment fecal calprotectin values were similar between the Aquamin treatment group and the placebo group (371 μg/gram versus 320 μg/gram). This included 8 of 14 placebo group values above the ≥160 μg/gram level that is considered a mark of ongoing inflammation (39) and 5 of 11 values in the 180-day Aquamin treatment group. At the end of treatment period, there was a 29.4% average decrease in fCAL values in the Aquamin treatment group while a 43.4% increase was seen in the placebo group. Among individuals receiving Aquamin for the 180-day period, 3 of 11 still had fecal-calprotectin values ≥160 μg/gram at the end of treatment period while 6 of 14 subjects in the placebo group were still above this “cut off” level (Figure 2D).

Blinded histological examination of H&E-stained sections of colonic biopsies was performed in parallel with fCAL measurements. Histological grading utilized a modified Geboes scoring (Figure 2E). The majority of pretreatment tissue sections were histologically normal or demonstrated only architectural evidence of past injury (abnormal crypt size and shape) (Figure 2E insert). Evidence of an inflammatory infiltrate was observed in five specimens (out of 28), and in three of these specimens, the inflammatory cells (mostly neutrophils) were present in the epithelium as well as in the lamina propria. Epithelial damage accompanying the inflammatory cell infiltration was observed in two of the three specimens. Given the lack of detrimental findings in most specimens, the average group scores were low (less than 1.0) to begin with. The group score increased among placebo recipients over the 90-day treatment period (120% increase), due primarily to the presence of the inflammatory infiltrate seen in four post-intervention specimens (Figure 2E and insert). In contrast, in the group of individuals receiving Aquamin for 180 days, the score decreased by 48%; this was due largely to reduced inflammatory cell infiltrate seen in the biopsy specimen from one of the two individuals with a high score at initiation (Figure 2E).

Figure 2F summarizes the pre- post-treatment findings for each parameter, comparing results from the Aquamin-treatment group and placebo. A composite index made up of the five individual parameters is also shown. The divergence in outcomes between the Aquamin-treatment and placebo groups is striking (p=0.0284 by unpaired t-test).

### Summary of findings after 90-days of Aquamin-treatment

At the end of the intervention period, there were a total of 28 subjects that had received Aquamin for at least 90-days (i.e., 12 subjects from the 180-day Aquamin treatment group at the intermediate time-point and 16 subjects from the placebo group that crossed over to Aquamin on day-90). To summarize, trends were similar to what was observed with Aquamin over the 180-day period but less definitive. For example, we observed at 3.3% decline in serum ALP, a 16% increase in ALPI and a 10.3% decline in CRP after 90 days of treatment with Aquamin. At the same time, there was a 18.9% increase in fCAL (compared to 43% increase in placebo). Histological analysis of colon specimens after 90 days of treatment revealed 41% decrease in the modified Geboes score as compared to 121% increase with placebo. Day-90 values for the entire group of subjects are presented in Supplement Figure 1.

### Subject perceptions (IBDQ scores and subject self-reporting)

The IBDQ questionnaire was employed as a way to assess subject perception of changes in their UC-related symptoms over the course of treatment. A score of 180 or higher (out of a total possible score of 224) on the complete questionnaire is considered to be indicative of the subject being in a state of remission. Consistent with the “in-remission” or mild disease classification, only 3 of the 28 enrolled subjects reported a score of <180 to begin with. Overall, there was little change in average group scores between day-0 and study completion. In the subjects consuming Aquamin for 180 days, average scores on the complete questionnaire improved from 200±13 to 204±13 while in the placebo group, scores at initiation and post-intervention were 199±20 and 200±18. In subjects who crossed over to Aquamin for 90 days, the IBDQ score increased from 200±18 to 202±14. With the 10-question, bowel-specific survey, pre- and post-intervention scores were 64±5 and 65±4, respectively in the Aquamin-treatment group while in the placebo group, post-intervention scores were 63±8 and 63±7, respectively.

At the end of their 180-day treatment period or at the 2-week post-intervention close-out call, subjects were given the opportunity to provide comments as to their perception of how the study had gone (Supplement Table 5). A total of 19 subjects (10 from the group that had received Aquamin for the complete 180-day period and 9 from those who were in the placebo group for the first 90 days and Aquamin for the final 90 days) chose to respond. To summarize, eleven subjects reported feeling better or having “more energy” and five subjects noted improvements in their gastrointestinal symptoms. Three subjects reported reduced usage of prescribed UC medicines and two reported decreased musculoskeletal pain (Supplement Table 5).

### Bone-related findings

Because bone loss is a common occurrence in UC (5) and because our earlier preclinical studies demonstrated inhibition of bone loss with Aquamin (23–25), each subject underwent DEXA scanning at initiation and at day-180. Shown in Figure 3 are BMD and BMC values obtained from two regions of interest – i.e., femoral neck and lumbar (L1-L4) vertebrae. Changes were seen in both regions but findings from the femoral neck region were of particular interest. Over the course of the six-month interventional period, the average femoral neck BMD value increased by 1.2% while the average BMC value increased by 3.4% (statistically significant compared to baseline) (Figure 3A). The increase in BMC relative to BMD reflects an increase in surface area captured by the DEXA scan (calculated to be 2.2% increase compared to pretreatment values). These improvements in bone structure assessed by DEXA scan were sufficient to drive a 7.3% (statistically significant) increase in hip bone strength calculated according to the method described by Yoshikawa et al (33) (Figure 3A). When BMD, BMC and area were assessed in the LI-L4 vertebrate, 1.4%, 2.1% and 1% increase were seen in subjects who received Aquamin for 180 days (Figure 3B). Representative DEXA scans from both regions of interest – upper femora and lumbar spine – carried out at Day 0 and Day 180 are shown in Figure 3C. In contrast to these findings, there was essentially no changes in BMD and BMC values at either site in subjects who were on placebo for the first 90 days and then crossed over to Aquamin for the last 90 days (Supplement Figure 2).

**Figure 3.**
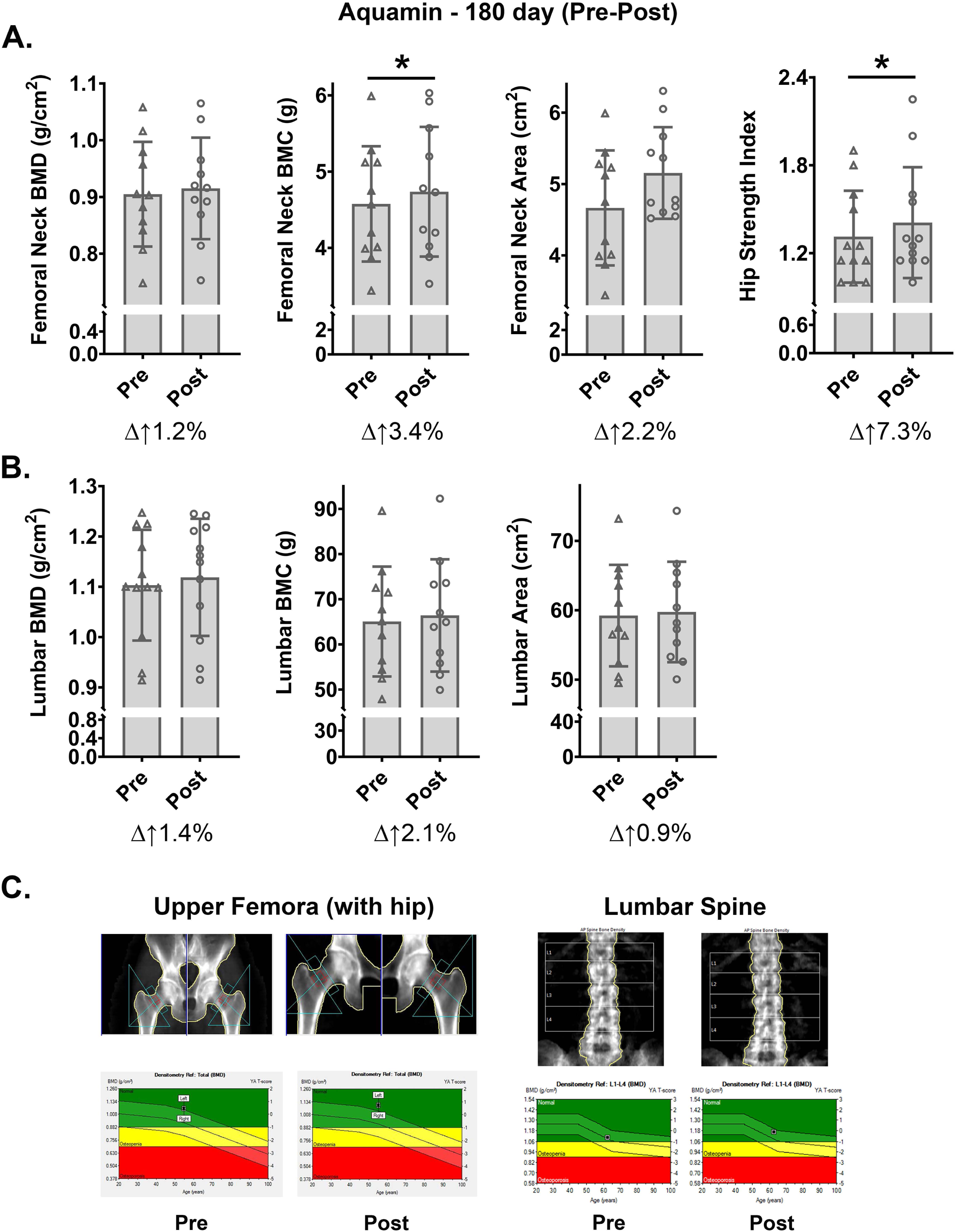
DEXA scan endpoints: Aquamin (180-day treatment group). **A**. Femoral neck. BMD, BMC and area values read directly from DEXA scans. Hip strength index was calculated according to Yoshikawa et al (33). **B**. Lumbar vertebrate. BMD, BMC and area values read directly from DEXA scans. All values are treatment group means and standard deviations. Delta values shown beneath each pair of bars indicate percentage change in the post-treatment value relative to the pretreatment value. Statistical significance between pre- and post-treatment values was determined by paired t-test. Asterisk (*) indicates P<0.05. **C**. Inserts: DEXA scans – upper femora (left) and lumbar (L1-L4) spine (right) performed at Day 0 (pre) and Day 180 (post). The graphs at the bottom of these scans demonstrate BMD values in both femora and spine. On the left, Aquamin improved femoral BMD by 3.4% in a 55-year-old patient, while on the right, lumbar BMD improved by 6% in a 63-year-old participant with 180 days of Aquamin intervention.

As part of the effort to understand bone changes observed in the DEXA scans, serum levels of osteocalcin and TRAP5b were measured by ELISA. Figures 4A and 4B demonstrate changes in both proteins. In subjects receiving Aquamin for 180 days, osteocalcin was increased by 34% and TRAP5b by 22%. In the placebo group, the average osteocalcin value was unchanged between pre- and post-treatment while the TRAP5b values decreased (22% reduction) (Figure 4A and 4B). In addition to osteocalcin and TRAP5b, we also assessed serum levels of bone-specific ALP (BALP). Consistent with previous findings (40), serum BALP accounted for a much greater percentage of total serum ALP than ALPI. Based on the average baseline values from all 28 subjects, BALP and ALPI levels were (170.4 ng/mL and 10.7 ng/mL, respectively). Among subjects receiving Aquamin for 180 days, BALP decreased by 9.4% while among subjects receiving placebo, BALP values increased by 6.7% (Figure 4C). Figure 4D summarizes the pre- post-treatment findings for each serum marker, comparing results from the Aquamin-treatment group and placebo. A composite index made up of the three independent markers is also shown. The divergence in outcomes between the Aquamin-treatment and placebo is striking (p=0.0136; unpaired t-test). Lastly, there was a decrease of 9% in serum osteocalcin, an increase of 4% in TRAP5b, and a decrease of 6% in BALP at the end of the study among individuals who began taking Aquamin at the day-90 timepoint (after crossing over). These trends were less convincing as compared to what was observed with Aquamin over the entire 180-day period but still improved over findings from the placebo group.

**Figure 4.**
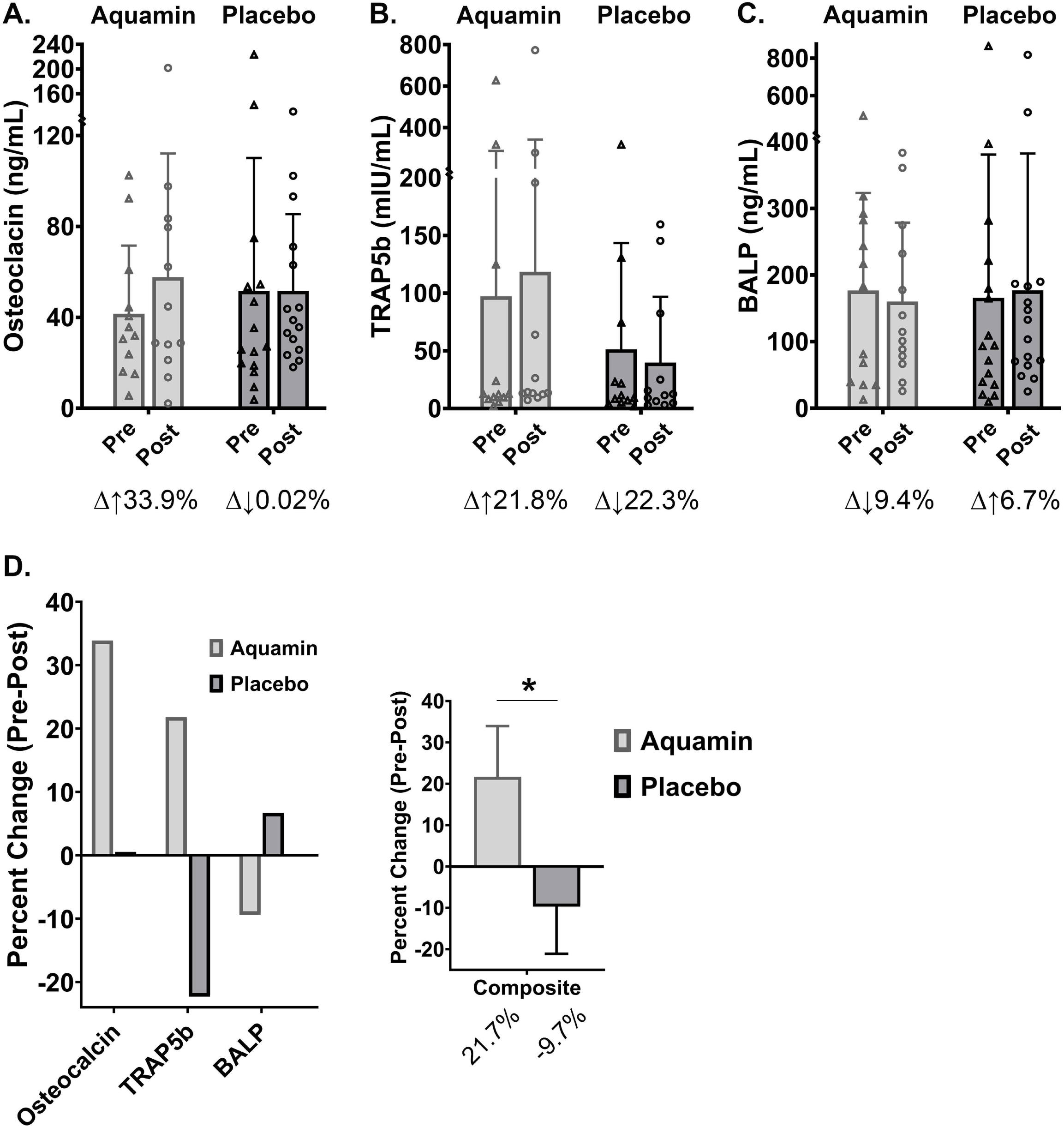
Bone-related chemistry endpoints: Aquamin (180-day treatment group) versus placebo. **A**. Serum osteocalcin: Values (ng/mL). **B**. Serum TRAP5b: Values (mIU/mL). **C.** Serum BALP: Values (ng/mL). All values determined by ELISA. Values are treatment group means and standard deviations. Delta values shown beneath each pair of bars indicate percentage change in the post-treatment value relative to the pretreatment value. **D.** Aquamin versus placebo comparison: Pre-post-treatment differences for each endpoint are plotted for the two participant groups to show the divergence between the groups. A composite score was generated by combining the three individual endpoint scores. Statistical significance between group (composite) values was determined by unpaired t-test. [Note: For composite graph on the right, inverse BALP values are plotted because while increased osteocalcin and TRAP5b are associated with improved bone metabolism, decreased BALP is reflective of improved bone metabolism.]

### Proteins of interest: proteomic analysis

Colonic biopsies obtained from each subject at day-0, day-90 and day-180 were examined using a data independent acquisition mass spectrometry approach to protein profiling as described in the Materials and Methods Section. The proteomic screen was used as a way to determine if treatment with Aquamin would have a detectable effect on proteins that contribute to epithelial cell differentiation and barrier formation. In parallel, proteins that are inflammation-related (including those that directly influence pathophysiology and those that may simply be reflective of an inflammatory environment) were also assessed as were moieties that contribute to electrolyte transport and fluid balance control in the colon. Findings from our previous preclinical studies with human colon tissue in organoid culture (17–21) and our earlier biomarker trial with healthy adults (22) were used to guide the present assessment. Protein expression data from this directed search are presented in Figure 5 as heatmap panels.

**Figure 5.**
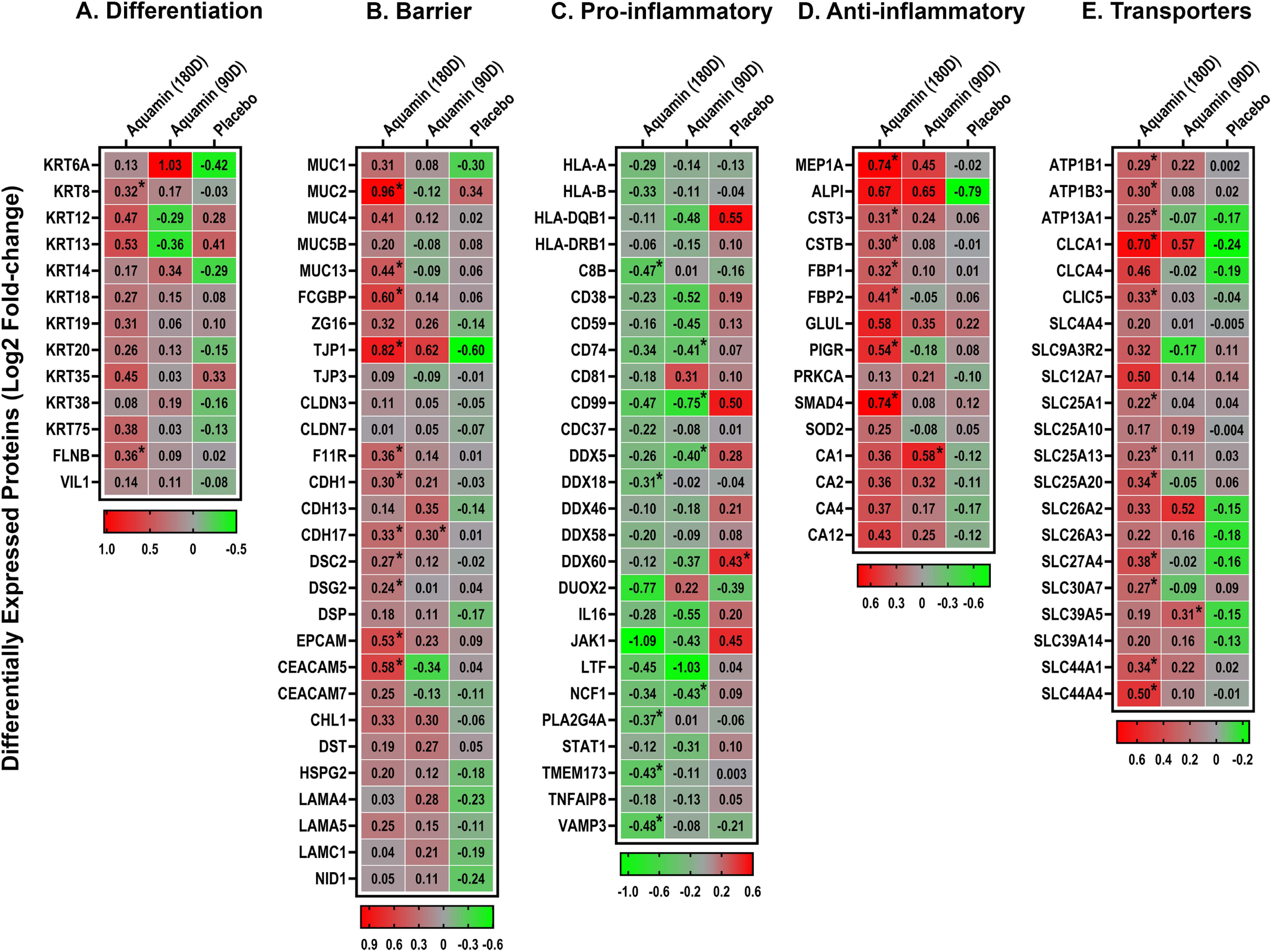
Proteins of interest based on directed search of the proteomic database. Proteins involved in A. differentiation, B. barrier formation, C &D. inflammation-related and E. transporters. Data are presented as a heatmap showing the pre- post-intervention changes (up- or down-regulation) of individual proteins of interest. Values reflect the Log2 abundance ratio from the respective pre-intervention levels from Placebo (n=16) and Aquamin treatment (n=12) for 180 days (180D). Protein expression is also shown from the subjects who were crossed over from placebo and ingested Aquamin for the last 90 days (90D) (n=16). Mean values for individual proteins were compared to respective pre-intervention values for statistical differences using the limma package. Asterisk (*) indicates a significant difference from the respective baseline expression at p<0.05.

Figure 5A and 5B demonstrate up-regulation of multiple proteins involved in differentiation (keratins) and barrier formation. Among barrier proteins, we saw enhanced expression of multiple cadherins, along with proteins involved in desmosome formation (i.e., DSC2, DSG2 and DSP). Tight junctional proteins, i.e., TJP1 (ZO1) and Junctional adhesion molecule A (JAM-A or F11R) were also strongly up-regulated but other components of tight junctions were only modestly induced or not detected. Both TJP1 and JAM-A play significant roles in tight junction formation and mucosal repair (41,42). Several mucins and other cell adhesion molecules that make up the mucinous layer were also elevated with intervention. Among these were MUC2 and FCGBP. These proteins are secreted by goblet cells; reduced expression is consistent with structural weakening of the mucus barrier, and this may occur prior to the onset of inflammation (43). Upregulation of these proteins with intervention is consistent, therefore, with improvement in the mucosal barrier structure. Another protein up-regulated with intervention (i.e., ZG16) suggests decreased binding of bacteria to the colonic epithelium (43,44). Additional adhesion molecules such as EPCAM and CHL1 (also known as L1CAM2) were also upregulated in response to Aquamin treatment, and their increased expression may have beneficial roles in immune homeostasis and barrier integrity (45,46). Finally, components of the basement membrane were responsive to Aquamin. These results are consistent with findings from our previous colon organoid culture studies (17,18,21). It is of interest to note that there was little change in expression levels for the majority of these same barrier proteins in the placebo group over the course of treatment (Figure 5B). Where pre- post-treatment differences were observed in the placebo group, the majority of the proteins were down-regulated post-treatment.

Figure 5C identifies a series of inflammation-related proteins that was down-regulated in response to Aquamin treatment. Although most of the moieties identified in this panel have multiple functions, several of these proteins are known to either directly contribute to inflammation or are part of the innate inflammatory response. For example, membrane-associated Phospholipase A2 (PLA2G2A) is frequently upregulated in gastrointestinal tract inflammation and is associated with epithelial cell damage (47); it was suppressed with Aquamin. Another potentially interesting example is the JAK1 protein; it was reduced by 2-fold with Aquamin while in the placebo group it was increased by 1.4-fold over the period of intervention. JAK1 is a tyrosine kinase and is critical for IFN-α/β signaling (48). Among other Aquamin-sensitive proteins was LTF, a neutrophil granule protein (49). The reduced level of this protein, like fecal calprotectin (Figure 2 above), is consistent with fewer neutrophils in the intestinal wall. Another Aquamin-down-regulated protein – i.e., neutrophil cytosol factor 1 (NCF1; also known as p47phox) – is required for activation of the NADPH oxidase as part of superoxide production in neutrophils (50). In summary, while proteomic profiling does not provide a detailed understanding of an individual protein’s role in gut inflammation, certain of the Aquamin-down-regulated proteins are likely to directly contribute to suppression of inflammatory pathophysiology. At the same time, the consistent down regulation of multiple proteins associated with the inflammatory state may simply be reflective of a lessening of inflammation.

Another group of inflammation-related proteins was up-regulated with Aquamin (Figure 5D). Proteins in this diverse group can play multiple roles in countering inflammation (e.g., have anti-protease or antioxidant activity, control antibody transport across cellular membranes, regulate cytokine generation) or may simply be a reflection of (in this case) a lessened inflammatory state. The up-regulation of SMAD4 is of particular interest in that deletion of this moiety is strongly correlated with bowel inflammation (51) as is PIGR, which signals through SMAD activation and plays a critical role in gut immune homeostasis. Its reduced expression has been noted in connection with the intestinal inflammation associated with IBD (45,52). MEP1A is another Aquamin-upregulated protein. Reduced MEP1A expression is correlated with increased disease activity (53). The finding of increased ALPI (in the biopsy specimens) is also of interest. An elevated level of ALPI in the tissue biopsies may help explain the reduction in the amount of enzyme shed into the plasma. In tissue, loss of ALPI function or reduced expression is associated with intestinal inflammation (54). Finally, the carbonic anhydrases are a family of enzymes that help maintain acid–base homeostasis, regulate pH, and fluid balance (55). Their decreased expression has been associated with UC (56); four isoenzymes were upregulated with Aquamin (Figure 5D).

Figure 5E shows expression levels for a number of integral membrane proteins that are responsible for establishing and maintaining electrochemical gradients. These proteins are essential for transport of a variety of organic solutes and inorganic minerals across plasma membranes and across intracellular membranes. As such, they play critical roles in osmoregulation in various tissues. In the colon, specifically, SLC26A3 (also known as DRA [down-regulated in adenoma]) plays a critical role in anion transport and fluid absorption (57). ATP1B1 and ATP1B3 also contribute to regulation of fluid absorption in the colon (58). What role (if any) that each of these Aquamin-up-regulated moieties presented in Figure 5E play in UC will require additional study.

It should be noted, finally, that we observed a high level of concordance between the two Aquamin cohorts in protein expression profiles; that is, the majority of proteins impacted by Aquamin at 180 days of treatment were also altered in the 90-day treatment cohort. The 90-day protein expression data are presented in the heatmap panels (Figure 5A-E) along with data from the other treatment groups.

Figure 6 identifies canonical pathways influenced by proteins shown in Figure 5. Supplement Table 6 provides the raw data upon which Figure 6 is constructed. These pathways (curated by Qiagen IPA) were identified based on the p-value of the overlap between identified proteins in the database (i.e., in Figure 5) and proteins in each given pathway (Figure 6A). Keratinization, Cell Junction Organization, Extracellular Matrix Organization, Transport of Inorganic Cations/Anions and Amino Acids/oligopeptides and Ion Channel Transport are among the top pathways identified in this manner. These are upregulated with Aquamin [with z-score greater than 2.2 and −log(p-value) of 2.75]. The z-score for the same pathways ranged from −0.28 to −2.3 with placebo treatment suggesting downregulation (Supplement Table 6). A key feature of Figure 6A is the strong dichotomy between the two interventions (Aquamin treatment for 180 days and placebo).

**Figure 6.**
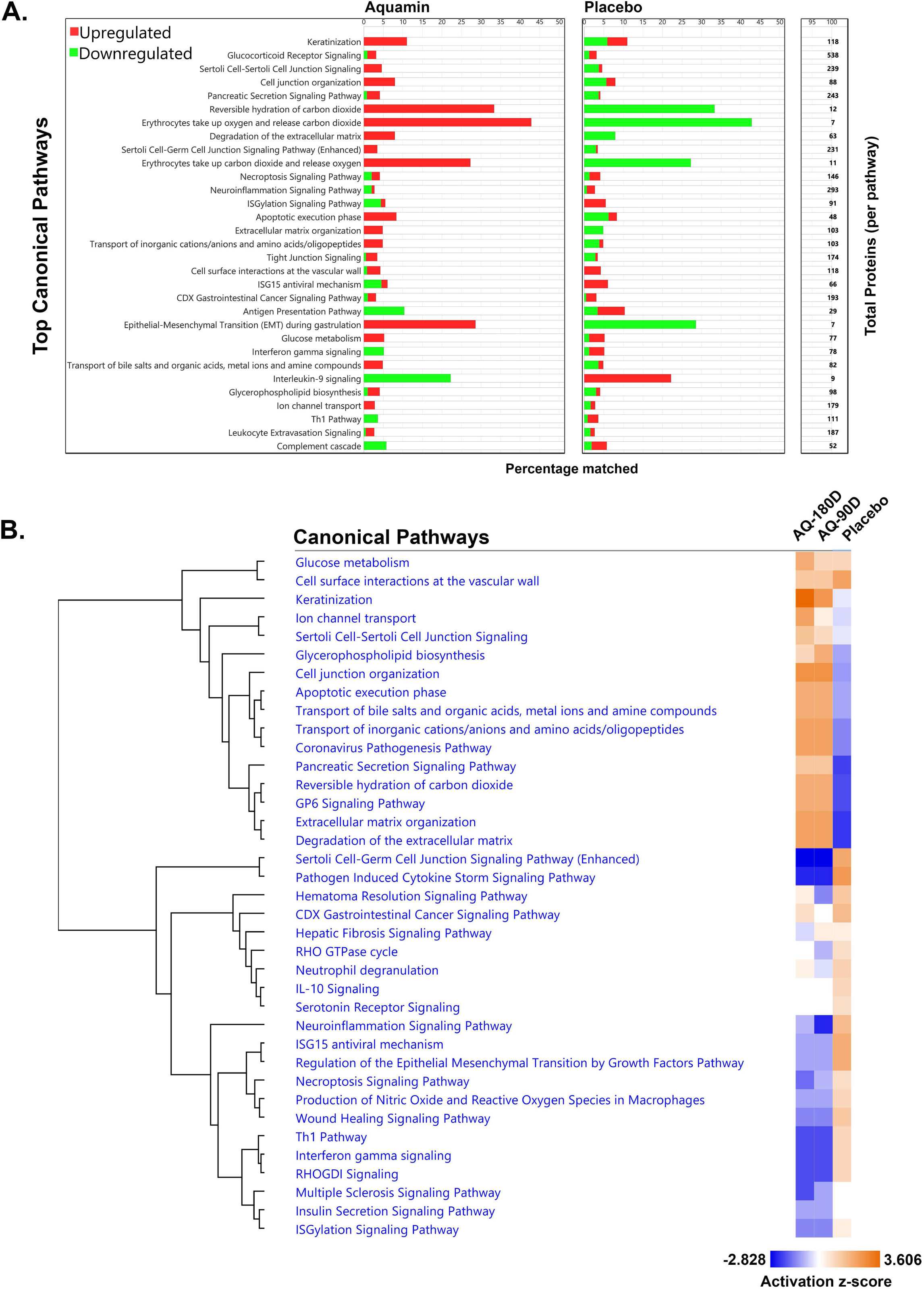
**A. Top Canonical Pathways**. Top canonical pathways affected (activated or suppressed) by the proteins shown in Figure 5. Pathways were curated by QIAGEN Ingenuity^®^ Pathway Analysis (IPA) and sorted based on the p-value. The stacked bar chart displays the percentage of proteins in the database compared to the total number of proteins contributing to the pathway. Up-regulated proteins (red), or down-regulated proteins (green) in each significantly enriched canonical Pathway are shown. The total number of proteins involved in each pathway is listed on the right margin of the graph. The left set of bars illustrates the pathways influenced by Aquamin (180-days)-responsive proteins, while the right set shows how the same pathways were affected by the placebo. **B. Comparative analysis of canonical pathways**. Pathways from the three treatment groups were compared using z-score activation and prioritized with hierarchical clusters and a heatmap for visualizing trends. Increased z-score activation is represented by orange color, while suppression is depicted by dark blue in the heatmap. The Aquamin groups (180- and 90-days) show concordance.

A comparative analysis of canonical pathways among all three treatment groups based on z-score activation is shown in Figure 6B. The key feature of this heatmap is the strong concordance between the two Aquamin-treatment groups and divergence of both with placebo. Some of the altered pathways listed include Pathogen Induced Cytokine Storm Signaling Pathway, Th1 Pathway, Interferon gamma signaling, Necroptosis Signaling Pathway, ISGylation Signaling Pathway, Neutrophil Extracellular Trap Signaling Pathway, Production of Nitric Oxide and Reactive Oxygen Species in Macrophages, ISG15 antiviral mechanism, Neuroinflammation Signaling Pathway, and IL-10 Signaling. These inflammation-related pathways were upregulated in placebo samples while Aquamin down-regulated the majority of pathways (Figure 6B). Supplement Table 6 provides the data from which this figure was constructed.

Additional analyses of the proteomic findings are presented in the Supplement. Supplement Figure 3 shows the distribution of proteins presented in Figure 5 by means of volcano plots. This figure demonstrates the extent of up- or down-regulation (abundance ratio) for each protein and the associated p-value for each protein in response to Aquamin (90- or 180-day) treatment or placebo. The divergence of the placebo group from the two Aquamin-treatment groups is apparent.

The Qiagen IPA analysis tool allows one to identify functional protein networks and to use the identified networks to predict involvement in biological functions and in disease processes. The top protein network identified in this manner (Gastrointestinal Disease, Inflammatory Response, Organismal Injury and Abnormalities) (Supplement Figure 4) reveals a network of proinflammatory molecules reduced in expression by Aquamin, while the same molecules are up-regulated in placebo. The raw data for this network analysis in provided in Supplement Table 7A-D. Supplement Table 7A identifies the seven top functional networks identified in this manner. Supplement Table 7B and 7C presents a list of molecules involved and relationships of their interactions in the top network. Supplement Table 7D presents disease or function annotations along with the predicted p-values for the molecules involved in these processes. Interestingly, colitis is listed as one of the top diseases with these altered molecules. Increased expression of CD74, NCF1, STING1 (TMEM173), and HLA-DQB1, as well as decreased expression of MEP1A, are the underlying causal molecules in placebo samples. Aquamin reversed their expression in virtually all instances.

## DISCUSSION

The present study is part of an effort to determine if (and to what extent) a multimineral intervention derived from marine red algae could have beneficial impact on individuals with UC. Preclinical studies have demonstrated a marked reduction in gastrointestinal inflammation when this intervention (Aquamin) is included in the diet over 15-18 months in healthy mice (13,14) or for 20 weeks in IL-10-/- mice (16). Studies with human colon tissue in organoid culture have demonstrated improved barrier structure/function with Aquamin (17–21). Finally, a 90-day pilot-phase biomarker trial carried out with healthy human subjects has demonstrated that many of the same changes seen in organoid culture can also be seen in human subjects with daily use of Aquamin (22,27). In the biomarker trial, no safety or tolerability issues were identified. Based on these past observations, we envision use of the multi-mineral product as an ancillary treatment in subjects with UC. As a step toward this goal, patients with UC in remission or those with mild disease were evaluated for changes in several disease-related biomarkers.

Over the 180-day treatment period, there were shifts in the levels of several individual biomarkers that, together, suggest the potential for disease improvement. Among these were reductions in serum ALP (both total and intestine-specific) as well as serum CRP and fecal calprotectin (markers of chronic inflammation and neutrophil accumulation in the colon [30,31], respectively). The reduction in fecal calprotectin was accompanied by fewer tissue neutrophils as observed in histological sections of colon biopsies. Pre- post-treatment differences in ALP reached a level of statistical significance, but while the improvements observed with CRP and colon neutrophil markers did not, improvements were not observed in the placebo group with any of these endpoints. This was clearly seen when a composite score based on the five biomarkers was generated for the Aquamin-treatment and placebo groups (Figure 2F).

The decrease in serum ALP was of particular interest in that of the eight subjects whose levels were above the upper normal reference range (100 U/L) value, the five subjects that were randomized to the placebo group showed no improvement over the course of treatment while the three subjects randomized to the Aquamin-treatment group all demonstrated reductions (12% average decrease). With serum ALPI in these subjects, the differences were event greater (27% decrease in the Aquamin-treatment group versus a 26% increase in placebo). Taken together, the biomarker findings allow us to suggest that even the low level of residual inflammation characteristic of our subject population may be amenable to further amelioration by multi-mineral intervention.

While the primary manifestations of UC reflect inflammation and superficial ulcer formation in the colonic wall, complications of the disease can be seen in other tissues. Bone loss is a common occurrence (5) and managing this is always a challenge (59). The pathophysiology of UC-related bone loss is not completely understood, but systemic inflammation and poor mineral absorption in the gut are both thought to be contributing factors (60). Given this understanding, subjects in our trial underwent DEXA scanning at the beginning of the study and at the day-180 timepoint. The response of the femur to Aquamin treatment suggests benefit to bone strength in two ways. First, the 3.4% increase in femoral neck BMC suggests increased bone accrual over the 6-month time frame. Second, the 2.2% increase in femoral neck area suggests new bone deposition on the periosteal surface. Both of these effects would be expected to increase bone strength. These two benefits together were sufficient to drive a 7.3% increase in the calculated hip bone strength. With pharmaceutical interventions, improvements of this magnitude in BMD and BMC values often require 2-3 years (61). Improvements in BMD and BMC (1.4% and 2.1%) in the lumbar spine (L1-L4) were also evident, but the ratio of BMC to BMD was not as striking as seen in the femoral neck. This was not unexpected since the relative amount of trabecular bone compared to cortical bone in the vertebrae is higher than in the femoral neck, making surface area expansion more difficult. Still, the greater increase in the BMC value compared to the BMD value is an indication of an increase in bone surface area and provides a rationale for increased strength (62).

Along with the changes seen by DEXA scan, we also observed increases in both osteocalcin (osteoblast marker; 34%) and TRAP5b (osteoclast marker; 22%) in serum Figures 4A and 4B). At the same time, bone-specific ALP (an indicator of high bone turnover in several conditions including UC [63]) decreased by 9% in the 180-day Aquamin cohort (Figure 4C). A composite score made up of the three bone markers demonstrated the divergence between Aquamin-treatment and placebo (Figure 4D). If similar results to those shown here can be observed in a larger trial, the benefit to bone health would be sufficient, we believe, to justify consideration for use in UC, regardless of how much improvement in colon-specific biomarkers were seen. The potential for reduction in bone loss independent of UC should also be considered.

Mechanism(s) contributing to bone improvement cannot be determined from the present work, but our previous studies in mice may provide insight. In the pre-clinical studies, dietary Aquamin supplementation reduced bone mineral loss and improved bone strength and stiffness (23–25). In those studies, there were changes in bone content of several cationic elements. The most striking change was a large (6-10-fold) increase in bone strontium. When available, strontium replaces some of the calcium in the bone lattice to produce a crystalline structure that is harder (64,65). Since strontium is one of the most prevalent cationic elements in Aquamin (Supplement Table 1), the beneficial activity of Aquamin on bone may reflect increased mineralization, *per se*. At the same time, previous work has shown that strontium stimulates the activity of both osteoblasts and osteoclasts to enhance bone turnover (66,67). Aquamin’s main action, therefore, may be mediated through effects on bone cell metabolism. While strontium [and calcium, of course (68)] are central to the beneficial effects of Aquamin on bone health, previous studies have suggested that other trace elements also contribute to bone health (69,70). These minerals are also present in Aquamin (Supplement Table 1). Finally, inflammation itself is a contributor to bone loss (59,60); thus, any reduction in systemic inflammation achieved by mineral supplementation (either directly or indirectly) could beneficially impact bone. Unfortunately, the experimental approaches needed to distinguish between these and other possibilities are not readily amenable to the type of study conducted here.

Liver involvement is another frequent occurrence in UC. Inflammation of the bile ducts with potential for scarring is a particular concern. Up to 80% of individuals diagnosed with primary biliary cholangitis (PBC) have underlying UC while up to 5% of individuals with UC have some degree of biliary tract inflammation (4,71,72). Often there is evidence of liver involvement before bowel symptoms are present. While elevated serum ALP can be an indication of inflammation in the wall of the gastrointestinal tract, elevated levels can also reflect concomitant bile duct injury or injury in the absence of detectable gastrointestinal wall inflammation. Unfortunately, while we were able to utilize sensitive ELISAs to distinguish intestine-derived and bone-derived ALP, there is no readily available way to identify and quantify hepatic contribution to the measured levels. Thus, there is no direct evidence from the current study for an Aquamin benefit in the liver. However, a recently published clinical study in patients with PBC, has utilized a reduction in serum ALP levels as a primary endpoint (73). Analogous to what was seen with bone, however, our long-term (15-18 month) studies in mice demonstrated beneficial effects in the liver with dietary Aquamin. This included a reduction in liver inflammation along with reduced fibrotic changes and an almost complete elimination of tumor formation (15). In a 20-week follow-up study in high fat diet-fed mice (74), Aquamin reduced the production of toxic lipid intermediates and oxidants in association with a reduction in liver inflammation seen histologically. Finally, in our trial with healthy subjects, Aquamin reduced the total bile acid content in the colon as well as levels of several toxic secondary bile acids (27). These past findings demonstrate that mineral supplementation can benefit the liver, but whether positive effects in the liver will be seen clinically requires further work.

As part of the present study, a proteomic screen was used to look for changes in a broad range of potentially relevant proteins. UC subjects who received Aquamin for 180 days demonstrated up-regulation of several proteins that contribute to the gastrointestinal barrier. In parallel, proteins associated with inflammation were down-regulated while several proteins with anti-inflammatory potential were up-regulated. Thus, the findings presented here are consistent with conclusions reached previously – i.e., that improved barrier structure and decreased inflammation are both consequences of multi-mineral intervention. Inflammation is, unarguably, a contributor to barrier breakdown in UC (1,2). However, barrier dysfunction has been noted in UC patients even in the absence of inflammation (43,75). Further, our own prior studies have demonstrated improved barrier structure / function in organoids from healthy subjects (17), and increased barrier protein expression in biopsies from healthy human subjects following Aquamin intervention (22). Based on this, we suggest that a multi-mineral intervention to improve the gastrointestinal barrier could complement therapeutic approaches that target inflammation directly.

Finally, both in the studies reported here and in our past studies (18,19) Aquamin treatment modulated expression of several cell surface transporter proteins, including moieties that help regulate fluid resorption in the colon. This activity could help to improve UC symptomatology even without affecting a major change in trans-epithelial barrier function and/or inflammation. While the present study did not address this issue directly, several of the subjects self-reported improvements in bowel function as well as an overall improvement in “well-being” at their final visit (see Supplement Table 5).

Taken together, the data from this study demonstrate that daily-ingestion of a marine algae-derived multi-mineral product has the potential to improve several biomarkers of gastrointestinal health (as well as bone health) in UC patients. While in our view these findings support continued research into the use of Aquamin as an ancillary intervention in UC, there were obvious limitations to the study. Most importantly, the number of subjects (twenty-eight in total) was small; the duration of treatment (180 days) was relatively short; the interventional agent was given at a low dose (normalized to provide 800 mg of calcium per day) and, most importantly, the subjects in the trial were from a cohort with UC in remission or at the mild stage. All of these deficiencies could be corrected with a larger trial. Our hope is to conduct a 1-2 year open-label trial, pending approval by FDA for such an effort. The lack of significant drug-related adverse events, the absence of negative changes in the comprehensive serum chemistry panel and the absence of tolerability issues should support approval for a longer-term study.

Finally, and perhaps, most importantly, a subsequent trial could include patients with moderate to severe UC. Because we envision the multi-mineral supplement to be used as an ancillary treatment, more seriously ill patients could remain on their primary therapy as dictated by standard of care while consuming the multi-mineral supplement. The mineral supplement might help not only to counteract pathophysiological events linked directly to inflammation and tissue damage but, with its potential for barrier-improvement, could retard rapid drug loss through the “leaky gut,” which is an issue in some patients with active UC (76,77). Either effect would be beneficial.

Another issue is the incomplete understanding of potential therapeutic mechanisms. While the current study and past work all support barrier improvement as a primary effect of Aquamin, there is no direct evidence at this point to substantiate this in UC patients. As a way to address this important issue, we have begun a study to measure gastrointestinal permeability using the urine lactulose-mannitol ratio approach (78) (clintrials.gov: NCT04855799). When complete, this study should go a long way toward establishing the relationship between the protein changes seen here and functional changes in the gut barrier.

In summary, pre-clinical studies have provided compelling evidence that daily ingestion of a multi-mineral product can improve barrier structure/function and reduce pro-inflammatory changes in the gastrointestinal tract. Consistent with this past body of evidence, the results presented here demonstrate that the same multi-mineral product has the potential to improve disease related biomarkers in patients with UC. Continued development as an ancillary treatment in UC is justified, we believe, by this work. We envision using Aquamin standardized to deliver no more than 1000 mg of calcium per day. Aquamin dosage standardized to deliver this amount of calcium would be within the range approved for long-term use as a dietary supplement. At this level, beneficial effects of calcium may be seen without unwanted side-effects (79). At the same time, the wide range of additional trace metals in the product could provide benefits not achievable with calcium alone (80). While, hopefully, benefits would not be limited to individuals with minimal disease, people with UC in remission or those with mild to moderate disease might benefit most from this approach. Current therapeutic development is focused mostly on addressing severe disease. Even though mild and moderate disease represents approximately 70% of the population with UC, there have been no therapeutic improvements to specifically address mild or less severe UC since the introduction of mesalamine in the 1980s (7). Identifying ways to impact mild to moderate disease is essential.

## Acknowledgement

The study team would like to thank the Michigan Institute for Clinical and Health Research (MICHR), the Michigan Clinical Research Unit (MCRU), the Nutrition, Exercise and Phenotype Testing Core for DEXA scanning, the Research Pharmacy, and the Clinical Trials Support Office at the University of Michigan. All of these groups provided invaluable support for the trial. Special thanks go to Mitch Seymour from the MICHR IND/IDE Investigator Assistance Program (MIAP) for his assistance with the IND process. Sherece Bank of the Clinical Research Management (CRM) team at Michigan also deserves our appreciation for her help with the RedCap database. We would also like to thank our study coordinators, Constantine Nolan and Almo Regazi, for their invaluable assistance with the study. The Histology Core and the Molecular Pathology Research Laboratory (MPRL) at the University of Michigan also deserve our gratitude for providing histology services. We would like to thank Marigot LTD (Cork, Ireland) for providing Aquamin capsules as a gift. Furthermore, we would like to acknowledge the IDeA National Resource for Quantitative Proteomics (NIH/NIGMS grant R24GM137786) for conducting proteomics analysis on a fee-for-service basis. Above all, the study would not have been possible without the participation of the study participants (patients with UC), and we are extremely thankful for their contribution.

## Data availability

The mass spectrometry proteomics dataset generated as part of this study are uploaded to a data repository called MassIVE (Mass Spectrometry Interactive Virtual Environment). The dataset will be freely accessible on the MassIVE repository (https://massive.ucsd.edu/v08/MSV000095472/) after the publication. (MassIVE and ProteomeXchange identifiers: MSV000095472 and PXD054344).

## SUPPLEMENT FILES

Supplement Text-File 1. Technical details of proteomic analysis.

Supplement Table 1. Aquamin Mineral Composition

Supplement Table 2. Disease Status and Drug Log

Supplement Table 3. Demographics

Supplement Table 4. Treatment-related reportable adverse events

Supplement Table 5. Subjective Feedback

Supplement Table 6. Canonical Pathways

Supplement Table 7. Functional networks identified by network analysis in Qiagen IPA

Supplement Figure 1. Chemistry and histology endpoints: Aquamin (90-day treatment group) versus placebo.

Supplement Figure 2. DEXA scan endpoints: Placebo / Aquamin (90-day treatment group).

Supplement Figure 3. Protein distribution by volcano plots.

Supplement Figure 4. IPA generated network 1

## SUPPLEMENT FIGURE LEGENDS

**Supplement Figure 1. Chemistry and histology endpoints: Aquamin (90-day treatment group) versus placebo. A**. Serum alkaline phosphatase (ALP): Values (U/L) determined as part of the serum chemistry panel. **B**. Serum intestine-specific alkaline phosphatase (ALPI): Values (ng/mL) determined by ELISA. **C**. C-reactive protein (CRP): Values (mg/dL) determined as part of serum chemistry panel. **D**. Fecal calprotectin (fCAL): Values (μg/g) determined by ELISA (BUHLMANN). **E**. Histological assessment: Values are presented as a modified Geboes score. All values are treatment group means and standard deviations. Delta values shown beneath each pair of bars indicate percentage change in the post-treatment value relative to the pretreatment value. **F.** Aquamin versus placebo comparison: Pre- post-treatment differences for each endpoint are plotted for the two participant groups to show the divergence between the two groups. A composite score was generated by combining the five individual endpoint scores.

**Supplement Figure 2. DEXA scan endpoints: Placebo / Aquamin (90-day treatment group).** As no DEXA was performed at the 90-day visit, this group represents subjects who were on placebo for the first 90 days and then on treatment after crossing over to Aquamin at day-90. **A**. Femoral neck. BMD, BMC, and area values read directly from DEXA scans. Hip strength index was calculated according to Yoshikawa et al (33). **B**. Lumbar vertebrate. BMD, BMC and area values read directly from DEXA scans. All values are placebo / Aquamin treatment group means and standard deviations. Delta values shown beneath each pair of bars indicate percentage change in the post-treatment value relative to the pretreatment value.

**Supplement Figure 3. Protein distribution.** Proteins presented in Figure 5 are shown by the volcano plots. **Top.** Aquamin treatment group (180 days), **Middle.** Aquamin treatment group (90 days), **Bottom.** Placebo. The abundance ratio (x-axis) for each protein is presented in Log2 transformed values and p-value (y-axis) is presented in −Log10 values. Red dots represent upregulated proteins, and green dots represent downregulated proteins.

**Supplement Figure 4. A subcellular layout of IPA generated network Analysis.** One of the top networks, shown in the figure – Network 1 – is associated with gastrointestinal disease, inflammatory response, and organismal injury. This network comprises 21 molecules from our dataset with a score of 39. Each network is limited to 35 molecules (IPA default setting) to ensure it is easy to follow. The likelihood of these molecules being part of the network is indicated by a p-value calculation. These networks were generated by proteins altered by Aquamin (**top**) and Placebo (**bottom**). The figure legend explains the confidence levels associated with the predicted activity and interaction among these molecules. These interactions are predicted based on the measured expression levels of the molecules. It is worth noting that the network molecular expression and interactions among these molecules in the Aquamin and placebo groups are completely opposite. The figure legends use color-coding to represent the expression and interactions with dark and light colors, respectively, to illustrate the predicted differences.

