## Supplementary material for "A MULTI-MINERAL INTERVENTION TO IMPROVE DISEASE-RELATED AND MECHANISTIC BIOMARKERS IN ULCERATIVE COLITIS PATIENTS": S Table 1

| **Supplement Table 1. Composition of Aquamin^®^ TG: Daily element intake** | | | | | |
| --- | --- | --- | --- | --- | --- |
| Element | μg/day | Element | μg/day | Element | μg/day |
| Aluminum | 189 | Hafnium | 0.3 | Rubidium | 0.06 |
| Antimony | <1.3 | Holmium | 0.1 | Ruthenium | 5.2 |
| Arsenic | 3.1 | Indium | <0.003 | Samarium | 0.3 |
| Barium | 19 | Iodine | 0.3 | Scandium | 4.4 |
| Beryllium | 4.6 | Iridium | <0.003 | Selenium | <1.3 |
| Bismuth | <1.3 | Iron | 1562 | Silicon | 233 |
| Boron | 88 | Lanthanum | 5.1 | Silver | 9.1 |
| Cadmium | 1.7 | Lead | 0.4 | Sodium | 12528 |
| Calcium | 800000 | Lithium | <1.3 | Strontium | 6033 |
| Carbon | 317949 | Lutetium | 0.1 | Sulfur | 8264 |
| Cerium | 2 | Magnesium | 66154 | Tantalum | 0.02 |
| Cesium | 0.005 | Manganese | 111 | Tellurium | <1.3 |
| Chloride | 5903 | Mercury | <0.003 | Terbium | 0.06 |
| Chromium | 7.7 | Molybdenum | <1.3 | Thallium | <1.3 |
| Cobalt | 4.8 | Neodymium | 1.2 | Thorium | 11 |
| Copper | 8.4 | Nickel | 4.6 | Thulium | 0.04 |
| Dysprosium | 0.4 | Niobium | 31 | Tin | 0.6 |
| Erbium | 0.3 | Osmium | 0.003 | Titanium | 82 |
| Europium | 0.1 | Palladium | 1.2 | Tungsten | <1.3 |
| Fluoride | 9 | Phosphorous | 200 | Vanadium | 9.4 |
| Gadolinium | 0.3 | Platinum | <0.003 | Ytterbium | 0.2 |
| Gallium | 0.3 | Potassium | 513 | Yttrium | 9.4 |
| Germanium | <0.003 | Praseodymium | 0.3 | Zinc | 7.8 |
| Gold | <1.3 | Rhenium | 0.01 | Zirconium | 14 |
|  |  | Rhodium | 1 |  |  |

The trace mineral composition of Aquamin^®^ TG was determined by an independent laboratory (Advanced Laboratories, Inc., Salt Lake City) for Marigot Limited (Ireland) in 2018.

Individual trace element composition was determined by Inductively Coupled Plasma Optical Emission Spectroscopy (ICP-OES) except for carbon (determined by ASTM D-1552), chloride and iodine (determined by Titration), and fluoride (determined by AOAC 939.11).

Values are presented based on daily consumption of Aquamin^®^ normalized to provide 800 mg of calcium.
