## Supplementary material for "A MULTI-MINERAL INTERVENTION TO IMPROVE DISEASE-RELATED AND MECHANISTIC BIOMARKERS IN ULCERATIVE COLITIS PATIENTS": S Table 3

| **Supplement Table 3. Demographics for subjects who completed 180-Day intervention** | | | | | | |
| --- | --- | --- | --- | --- | --- | --- |
| **A. Study cohort demographics** |  |  |  |  |  |  |
|  | **Female** | | **Male** | | **Both Genders** | |
| **Ethnic Category** | N | % | N | % | Total | % |
| Hispanic or Latino | 0 | 0.0% | 0 | 0.0% | 0 | 0.0% |
| Not Hispanic or Latino | 16 | 57.1% | 11 | 39.3% | 27 | 96.4% |
| Unknown | 0 | 0.0% | 0 | 0.0% | 0 | 0.0% |
| Prefer not to answer | 1 | 3.6% | 0 | 0.0% | 1 | 3.6% |
| **Total** | **17** | **60.7%** | **11** | **39.3%** | **28** | **100.0%** |
| **Racial Category** *(single category per participant)* | N | % | N | % | Total | % |
| American Indian/Alaska Native | 0 | 0.0% | 0 | 0.0% | 0 | 0.0% |
| Asian | 0 | 0.0% | 0 | 0.0% | 0 | 0.0% |
| Native Hawaiian or Other Pacific Islander | 0 | 0.0% | 0 | 0.0% | 0 | 0.0% |
| Black or African American | 1 | 3.6% | 0 | 0.0% | 1 | 3.6% |
| White | 16 | 57.1% | 10 | 35.7% | 26 | 92.9% |
| Other | 0 | 0.0% | 1 | 3.6% | 1 | 3.6% |
| Unknown | 0 | 0.0% | 0 | 0.0% | 0 | 0.0% |
| **Total** | **17** | **60.7%** | **11** | **39.3%** | **28** | **100.0%** |
| **Age at Enrollment Category** | N | % | N | % | Total | % |
| 18 - 21 years | 1 | 3.6% | 1 | 3.6% | 2 | 7.1% |
| 22 - 29 years | 3 | 10.7% | 3 | 10.7% | 6 | 21.4% |
| 30 - 39 years | 2 | 7.1% | 4 | 14.3% | 6 | 21.4% |
| 40 - 49 years | 4 | 14.3% | 1 | 3.6% | 5 | 17.9% |
| 50 - 59 years | 2 | 7.1% | 0 | 0.0% | 2 | 7.1% |
| 60 - 69 years | 3 | 10.7% | 2 | 7.1% | 5 | 17.9% |
| 70 - 79 years | 2 | 7.1% | 0 | 0.0% | 2 | 7.1% |
| > 80 years | 0 | 0.0% | 0 | 0.0% | 0 | 0.0% |
| **Total** | **17** | **60.7%** | **11** | **39.3%** | **28** | **100.0%** |
| **B. Gender and Age:** |  |  |  |  |  |  |
|  | Gender | Age (Y) |  |  |  |  |
| Placebo (n=16) | M:6 / F:10 | 43.1±15 |  |  |  |  |
| Aquamin (n=12) | M:5 / F:7 | 50.1±18 |  |  |  |  |
| Age presented in years at the start. |  |  |  |  |  |  |
| **C. Body Mass Index:** | BMI - Weight/Height^2^ (Kg/m^2^) | | |  |  |  |
|  | Pre | Post |  |  |  |  |
| Placebo | 25.3±5.1 | 25.5±5.2 |  |  |  |  |
| Aquamin | 26.8±4.7 | 27.1±4.5 |  |  |  |  |
| **D. Calcium Intake Levels at the start (Estimated by DHQ3)** | | | | | | |
|  | Calcium (mg/day) | |  |  |  |  |
| Placebo (P) | 1134±486 |  |  |  |  |  |
| Aquamin (AQ) | 1083±439 |  |  |  |  |  |
| Subjects above 1000 mg/day Calcium: 8 (Placebo / AQ-90d) and 7 (AQ-180d) | | | | | | |
