## Supplementary material for "A MULTI-MINERAL INTERVENTION TO IMPROVE DISEASE-RELATED AND MECHANISTIC BIOMARKERS IN ULCERATIVE COLITIS PATIENTS": S Table 4

| **Supplement Table 4. Treatment-Related Reportable Adverse Events** | | | |
| --- | --- | --- | --- |
| **Event** | ***Placebo**** | ***AQ-90d**** | **AQ-180d** |
| Number of subjects participated | 16 | 16 | 12 |
| Number of subjects reporting events | 14 | 7 | 12 |
| Total number of adverse events | 39 | 14 | 44 |
| COVID-19 infection | 0 | 1 (1) | 3 (3) |
| Sore throat (with enlarged lymph nodes) | 0 | 1 (1) | 1 (1)^a^ |
| Earache | 0 | 0 | 1 (1)^a^ |
| Neuropathy | 1 (1) | 0 | 0 |
| Anemia (low iron) | 0 | 0 | 1 (1) |
| Elevated blood pressure | 1 (1) | 0 | 1 (1) |
| Dizziness (light headedness) | 2 (2)^b^ | 0 | 0 |
| Headache | 3 (3) | 1 (1) | 1 (1) |
| Bilateral flank pain | 1 (1) | 0 | 0 |
| Uterine Polyp (with bleeding) | 1 (1) | 0 | 0 |
| Fatigue | 1 (1) | 1 (1) | 2 (2)^c^ |
| Back pain | 0 | 1 (1) | 0 |
| Joint pain | 0 | 0 | 5 (1) |
| Physical injury (car accident) | 0 | 1 (1) | 0 |
| Tooth infection | 0 | 0 | 1 (1) |
| Tooth removal | 0 | 1 (1) | 0 |
| Jaw discomfort (with increased clenching) | 0 | 1 (1) | 0 |
| Gastrointestinal events |  |  |  |
| *Nausea* | 3 (2) | 0 | 0 |
| *Flatulence (& bloating)* | 20 (9) | 1 (1) | 21 (9) |
| *Abdominal Pain (gastric discomfort)* | 4 (4) | 1 (1) | 3 (3)^d^ |
| *Constipation* | 0 | 2 (1) | 0 |
| *Diarrhea (loose stool)* | 1 (1) | 0 | 0 |
| *Blood in stool* | 1 (1) | 2 (2) | 2 (1) |
| *UC flare* | 0 | 0 | 1 (1)^e^ |
| *Food poisoning* | 0 | 0 | 1 (1) |

*Number in the parenthesis represent subjects experiencing an adverse event (AE).*

**The 16 subjects in the placebo group and the 16 subjects in the *AQ-90d group are the same individuals. These subjects crossed over to Aquamin^®^ after 90 days. Adverse events listed under placebo were reported during the first 90 days and adverse events listed under AQ-90d were reported during the final 90 days of intervention. The adverse events reported by the 12 subjects in the AQ-180d group were reported over the entire treatment period.*

*^a^Associated with Covid-19 infection in one subject*

*^b^Could be due to medication (Saxenda) in one patient*

*^c^Associated with Covid-19 infection in two subjects*

*^d^One of the subjects reporting abdominal discomfort attributed this to being off of his regular UC treatment for 5 weeks due to lack of insurance*

*^e^ The flare was detected during the 90-day visit with improvement by the 180-day visit*
