## Supplementary material for "A MULTI-MINERAL INTERVENTION TO IMPROVE DISEASE-RELATED AND MECHANISTIC BIOMARKERS IN ULCERATIVE COLITIS PATIENTS": S Table 5

| **Supplement Table 5. Subjective Feedback: Response to Aquamin^®^** | |
| --- | --- |
| **Subject ID (Duration)** | **Participants reported or noted:** |
| Subject 1 (180-d) | feeling better, biologics (Tofacitinib) dose reduced by the GI doc after the study completion |
| Subject 2 (180-d) | improved energy level - improved - felt fatigued again 2 weeks after completing study |
| Subject 3 (180-d) | improved energy level - Improved. However, felt fatigued again after discontinuing Aquamin |
| Subject 4 (180-d) | feeling much better and felt comfortable; otherwise felt that flare could start anytime |
| Subject 5 (180-d) | improved stool consistency and improved frequency |
| Subject 6 (180-d) | hairs got better, Participant reported feeling much healthier |
| Subject 7 (180-d) | feeling good, Participant noted being without UC maintenance therapy due to lack of insurance |
| Subject 8 (180-d) | "90% on the way to remission” using own words, did complain of some blood in stool |
| Subject 9 (180-d) | feeling better in general and had better mobility with decreased arthritis issues. |
| Subject 10 (180-d) | commented, "I loved the treatment and felt great during my participation" |
| Subject 11 (90-d) | feeling better |
| Subject 12 (90-d) | feeling fatigued after completing study and noted that joint pains started again |
| Subject 13 (90-d) | that stool firmed up; Participant noted never having had that consistency of stool in last 20 years |
| Subject 14 (90-d) | firmer stool compared to pre-participation stage and less reactive to previous dietary triggers |
| Subject 15 (90-d) | feeling much better; Participant noted a decreased number of days with GI discomfort |
| Subject 16 (90-d) | feeling energetic and feeling much better as compared to last visit |
| Subject 17 (90-d) | patient experienced a flare 2 weeks after discontinuing Aquamin, which required a mesalamine suppository |
| Subject 18 (90-d) | improved stool consistency |
| Subject 19 (90-d) | mesalamine reduced to half-dose; decreased dysplastic changes in the colon (on GI surveillance) |

*Nineteen subjects provided additional feedback at the last study visit or at the close-out call two weeks later. For public dissemination, subject identifiers replaced by numbers 1 through 19.*

*180-d: Aquamin given for 180 days (n=12 subjects). 90-d: Aquamin given for last 90 days (n=16 subjects).*
