## Supplementary figures and images for "A MULTI-MINERAL INTERVENTION TO IMPROVE DISEASE-RELATED AND MECHANISTIC BIOMARKERS IN ULCERATIVE COLITIS PATIENTS"

### S Fig 1

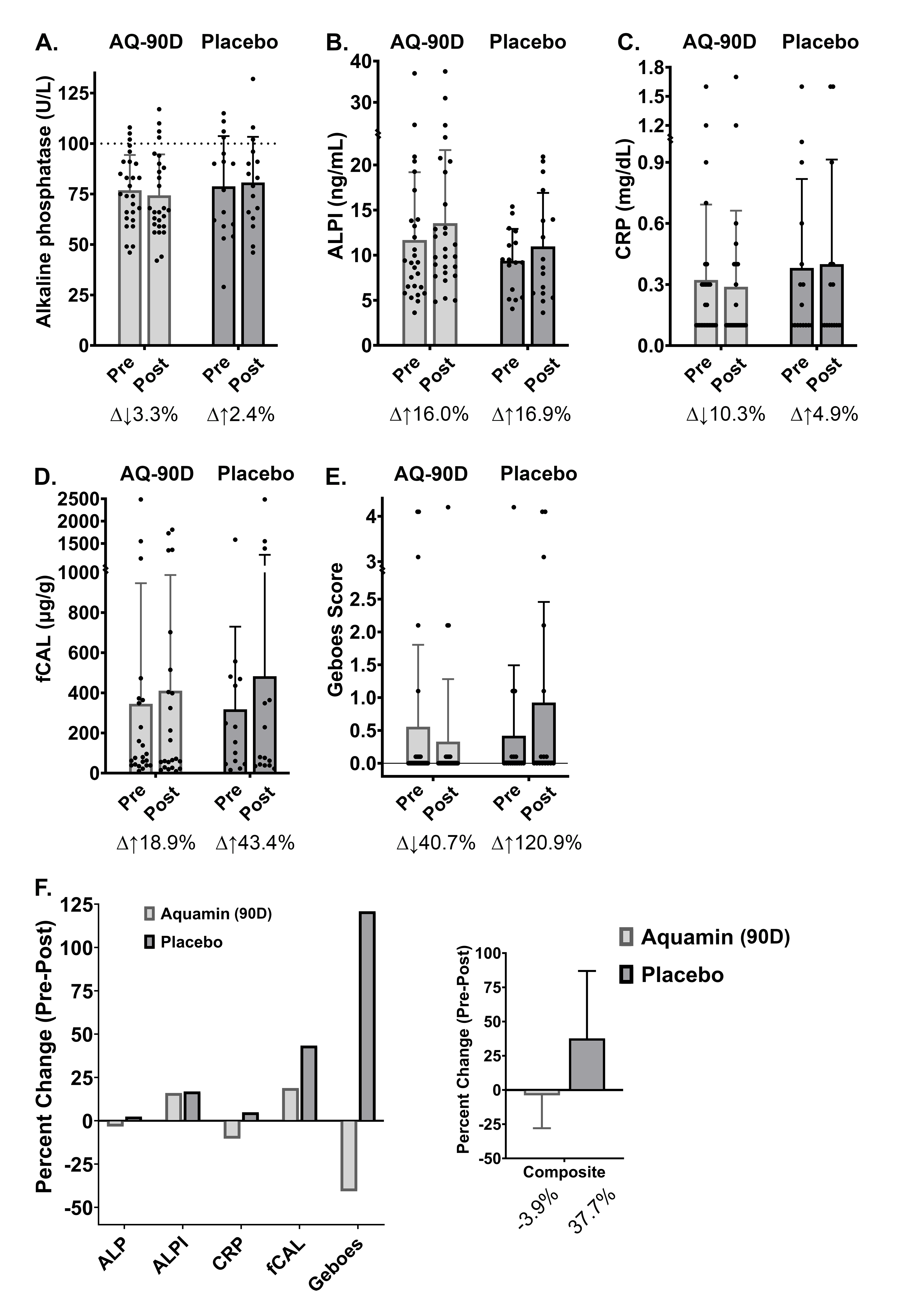

### S Fig 2

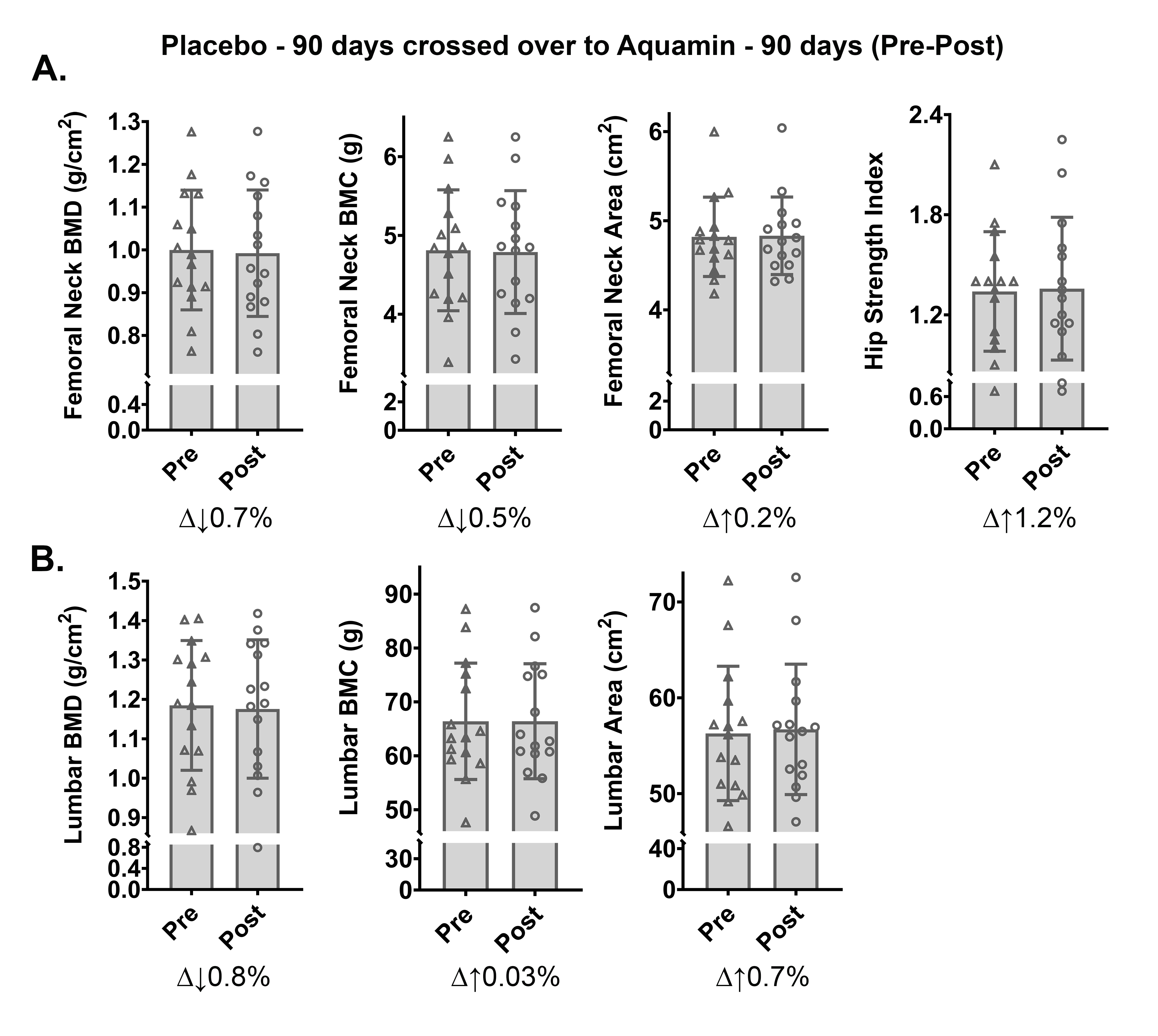

### S Fig 3

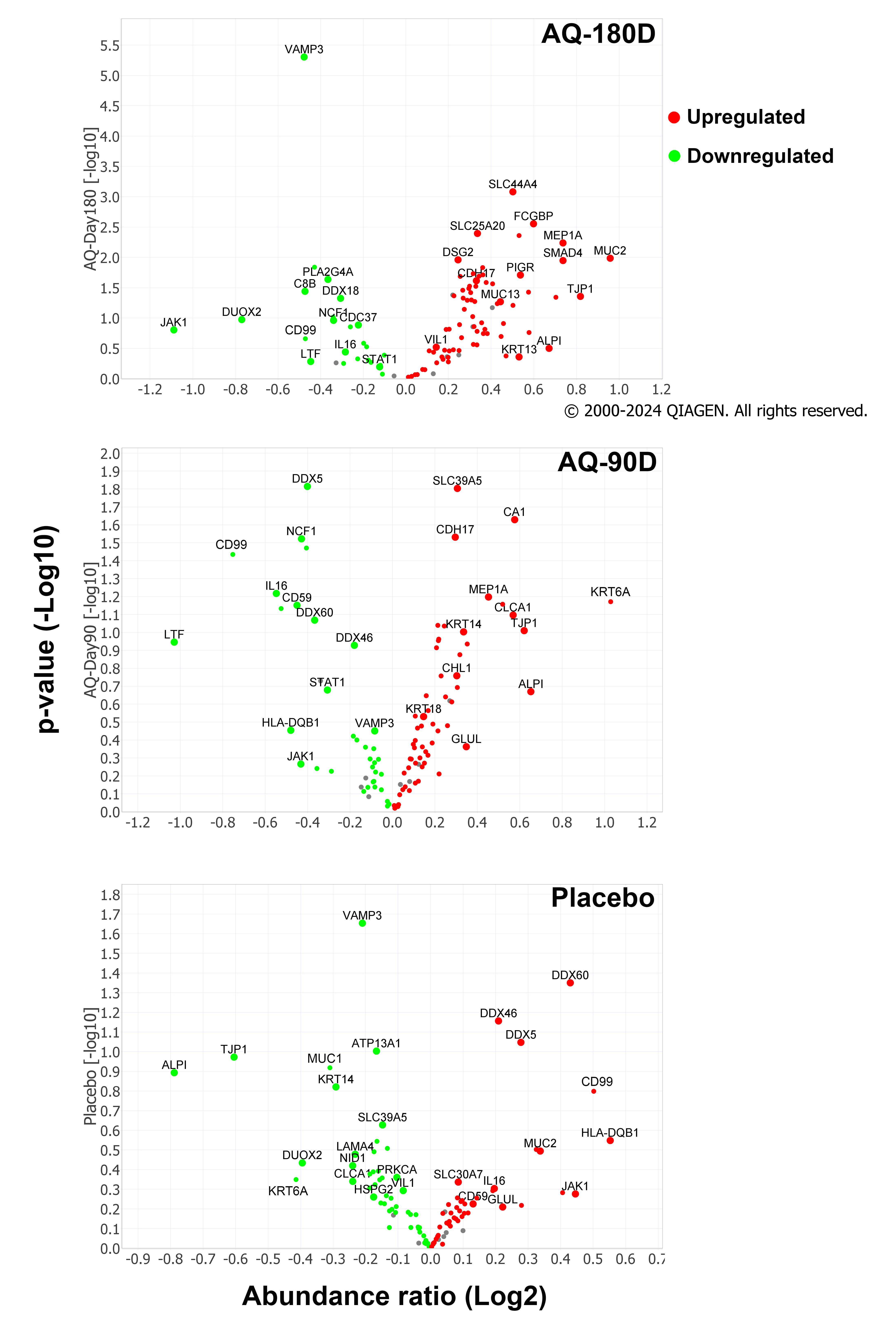

### S Fig 4

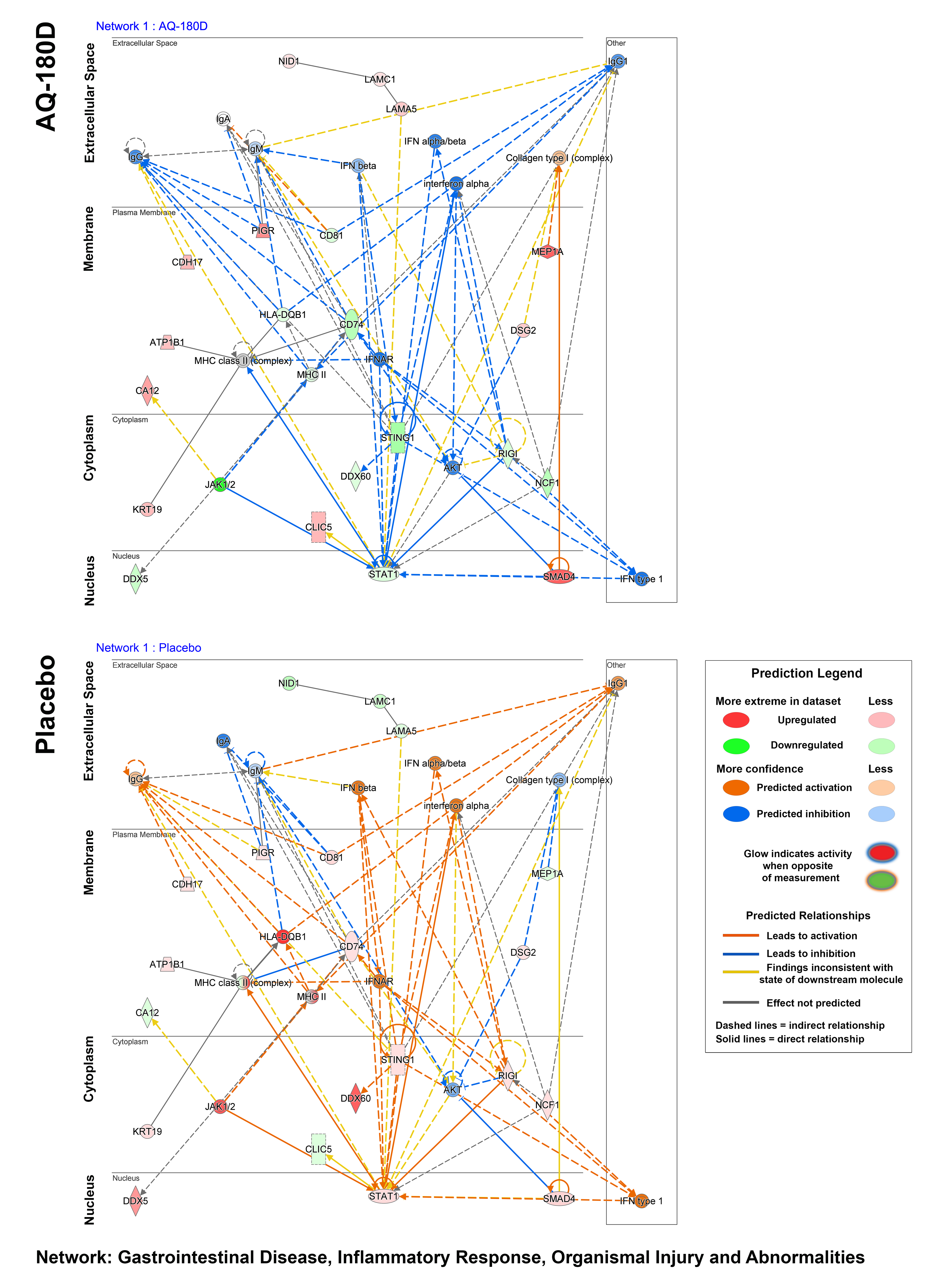
